# The epidemiological impact of digital and manual contact tracing on the SARS-CoV-2 epidemic in the Netherlands: empirical evidence

**DOI:** 10.1101/2023.04.27.23289149

**Authors:** Wianne Ter Haar, Jizzo Bodriesz, Roderick P. Venekamp, Ewoud Schuit, Susan van den Hof, Wolfgang Ebbers, Mirjam Kretzschmar, Jan Kluytmans, Carl Moons, Maarten Schim van der Loeff, Amy Matser, Janneke H. H. M. van de Wijgert

## Abstract

**Background:** The Dutch government introduced the CoronaMelder smartphone application for digital contact tracing (DCT) to complement manual contact tracing (MCT) by Public Health Services (PHS) during the 2020-2022 SARS-CoV-2 epidemic. Modelling studies showed great potential but empirical evidence of DCT and MCT impact is scarce.

**Methods:** We determined reasons for testing, and mean exposure-testing intervals by reason for testing, using routine data from PHS Amsterdam (1 December 2020 to 31 May 2021) and data from two SARS-CoV-2 rapid diagnostic test accuracy studies at other PHS sites in the Netherlands (14 December 2020 to 18 June 2021). Throughout the study periods, notification of DCT-identified contacts was via PHS contact-tracers, and self-testing was not yet widely available.

**Results:** The most commonly reported reason for testing was having symptoms. In asymptomatic individuals, it was having been warned by an index case. Only around 2% and 2-5% of all tests took place after DCT or MCT notification, respectively. About 20-36% of those who had received a DCT or MCT notification had symptoms at the time of test request. Test positivity after a DCT notification was significantly lower, and exposure-test intervals after a DCT or MCT notification were longer, than for the above-mentioned other reasons for testing.

**Conclusions:** Our data suggest that the impact of DCT and MCT on the SARS-CoV-2 epidemic in the Netherlands was limited. However, DCT impact might be enlarged if app use coverage is improved, contact-tracers are eliminated from the digital notification process to minimise delays, and DCT is combined with self-testing.

**Author summary:** During the 2020-2022 SARS-CoV-2 epidemic, the Dutch government introduced digital contact tracing (DCT) using a smartphone application to complement manual contact tracing (MCT) by professional contact-tracers. Mathematical models had suggested that DCT could slow down virus spread by identifying more individuals with whom the smartphone user had been in close contact and by reducing notification and testing delays after exposure. We used data collected during the Dutch epidemic to evaluate whether this was indeed the case and found that DCT and MCT had limited impact. Only around 2% of all tests took place after a DCT notification, and 2-5% after a MCT notification depending on MCT capacity at the time. Test positivity was lower after a DCT notification, and exposure-test intervals were longer after a DCT or MCT notification, than for other reasons for testing. About 20-36% of those who had received a DCT or MCT notification had symptoms at the time of test request and might have tested anyway even without having received the notification. However, DCT impact might be enlarged in future epidemics if app use coverage is improved and all exposure-notification-testing delays are minimised (e.g. no involvement of professional contact tracers and enabling self-testing after DCT notification).

## Introduction

Source and contact tracing by public health professionals (manual contact tracing or MCT) is a well-known method to control the spread of communicable diseases, and was used during the 2020-2022 SARS-CoV-2 pandemic. However, for the first time, digital contact tracing (DCT) using smartphone applications was also introduced in many countries worldwide (1–3). These apps differed in their levels of privacy-by-design, functionalities, and management of detected exposures. But they all had in common that a notification was triggered if the app user had been exposed to a person who had tested SARS-CoV-2-positive. In the Netherlands, the app was dubbed CoronaMelder. It was launched in the 8th month of the Dutch epidemic on 10 October 2020 (see *Supplementary introduction* for a detailed history of the Dutch epidemic) (4,5).

DCT has several theoretical advantages over MCT and over testing because of having symptoms or having received a warning by an index case. These include identification and notification of more contacts (including contacts whom the app user does not know personally), reducing delays in becoming aware of an exposure or presymptomatic infection and getting tested, and increasing general awareness which might stimulate preventive behaviour (6). Modelling studies from various countries in the first year of the pandemic suggested that DCT might have a prominent epidemiological impact, but that the impact depends on user uptake, use coverage, percentage of exposures detected, testing capacity and policies, and the specifics and delays of the tracing process (7–10). More empirical evidence is needed to properly quantify these factors for use in future mathematical transmission models, which are needed to guide policy decisions regarding future investments in DCT and MCT (1,2). In this paper, we assessed whether DCT and MCT had added value in SARS-CoV-2 control during the 2020-2021 SARS-CoV-2 epidemic in the Netherlands. We focused on reasons for testing (including having received an exposure notification via DCT or MCT) and exposure-test intervals by reason for testing.

## Materials and Methods

In the Netherlands, the general public could get tested at public health service (PHS) test sites (from 1 June 2020 onwards), commercial test sites (between February 2021 and March 2022), and by self-testing (from 31 March 2021 onwards). Public testing was free-of-charge for specific indications, and initially included symptomatic individuals only. Free-of-charge testing of exposed but asymptomatic individuals with close contacts became possible from 1 December 2020 onwards: the initial recommendation was to test on the fifth day after exposure, but from 18 February 2021 onwards to also test as soon as possible after the exposure. Individuals who tested positive at public or commercial test sites were contacted by a PHS employee and asked about their recent contacts; until February 2022, individuals with a positive self-test were asked to do a repeat test at a PHS test site to confirm the result and to enter the PHS MCT programme. During times when the MCT programme was fully operational, PHS employees notified each reported close contact. However, MCT was regularly scaled down due to the programme being overwhelmed, and at those times, cases were asked to notify their close contacts themselves (*Supplementary methods*).

The CoronaMelder app was introduced on 10 October 2020 (technical explanations in the *Supplementary introduction* and Figure S1). By the end of May 2021, almost 30% of the Dutch population had downloaded the CoronaMelder app but only around 18% were still actively using it (5). For DCT to have epidemiological impact, all steps of the exposure-notification-action cascade should be optimised (Figure 1 and *Supplementary introduction*): app uptake and active use, detection of exposures, quarantine and testing once an exposure is detected, and isolation and notification of contacts after testing positive. App users who tested positive could voluntarily upload that information (via a so-called PHS-key) from the app to a backend server, which subsequently resulted in notifications being sent to all phones with CoronaMelder in active mode that had been within 1.5 meters for at least 15 minutes within the last 14 days. PHS-keys could initially only be uploaded to the backend server after a PHS employee had entered the date of symptom onset or, if no symptoms present, the date of the positive test into the PHS-key. From 11 November 2021 onwards (which is after the three datasets used in this study had already been collected), app users could upload their PHS-key themselves.

**Figure 1:**
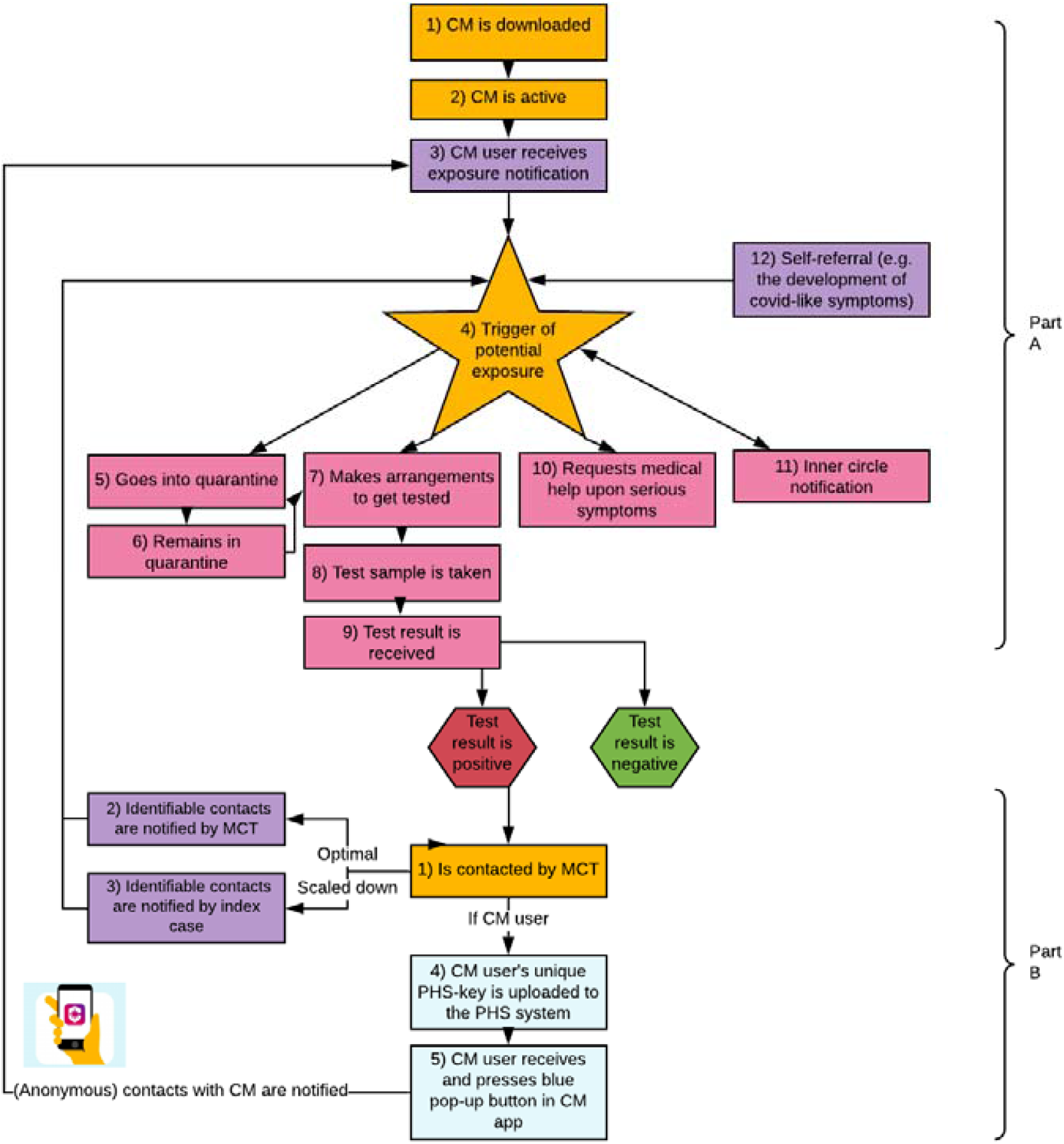
The exposure-notification-action cascade during the SARS-CoV-2 epidemic in the Netherlands, 2020-2021. See *Supplementary introduction* for detailed explanations of each of these steps.

### Data sources

We could not use data collected via CoronaMelder itself due to the privacy sensitive configuration of the app. Instead, we used routinely collected public health data at the PHS Amsterdam and data from two SARS-CoV-2 rapid diagnostic test (RDT) accuracy studies that were conducted at various other PHS test sites in the Netherlands.

PHS Amsterdam data were extracted from the national CoronIT, HPZone, and Osiris databases (described in the *Supplementary methods*). We plotted reasons for testing over time for the period 1 June 2020 (start of public testing for symptomatic individuals) until 31 May 2021, but conducted all analyses on data collected between 1 December 2020 (DCT available, and DCT and MCT-notified asymptomatic close contacts could also access public testing) and 31 May 2021. The PHS Amsterdam dataset contained CoronIT data between 1 December 2020 and 31 May 2021 on age, gender, postal code, reason for testing, symptoms at the time of test request, date/time of test appointment, date/time of testing, and test result for 562,159 tests (n_t_) by 372,545 individuals (n_i_) (Figure S2). Exposure-testing intervals could only be determined for individuals who had been part of the PHS Amsterdam MCT programme as either a case or a contact (the PHS Amsterdam MCT subset). CoronIT data were merged with exposure date data from HPZone (available up to 31 March 2021) and exposure level data from Osiris. After merging, this PHS Amsterdam MCT subset included 20,647 exposure-testing intervals (n_e-t_) by 20,355 individuals (n_i_) (Figure S2; merging process described in the *Supplementary methods*).

The first RDT study was conducted between 14 December 2020 and 6 February 2021 at PHS test sites in the West-Brabant (Raamsdonksveer and Roosendaal) and the Rotterdam-Rijnmond (Rotterdam The Hague Airport and Ahoy) regions. The second RDT study took place between 12 April and 18 June 2021 at PHS test sites in the West-Brabant (Breda), Rotterdam-Rijnmond (Rotterdam The Hague Airport and Ahoy), and IJsselland (Zwolle) regions. The primary diagnostic accuracy results have been published elsewhere (11,12). Participants provided written informed consent and had to be at least 16 years. In the first RDT study, close contacts who were asymptomatic at the time of test request were eligible for participation, but about 9% of them had developed symptoms by the time of testing a few days later. The second RDT study included all individuals who scheduled a SARS-CoV-2 test at the participating PHS sites irrespective of reason for testing. In both studies, participants were asked to complete short questionnaires developed by the study teams – including questions about reasons for testing and CoronaMelder use – while waiting to be sampled at the test site (Figure S3). The questionnaire data were merged with the PHS test data including test results. The first RDT study dataset contained 4,126 individuals (Figure S4). The second RDT study dataset contained 7,925 individuals but date of last exposure, and thereby an exposure-testing interval, was only available for 3,172 individuals (Figure S4).

### Statistical analyses

In all datasets, individuals could report multiple reasons for testing (Table 1 for an overview of these reasons). Symptoms could have been reported as a reason for testing, but anyone requesting a test or participating in one of the studies was also asked if they had any symptoms as a standalone question. These questions were asked at the time of test request in the PHS Amsterdam dataset and at the time of testing in the two RDT studies. In most analyses (unless noted otherwise below), we used a hierarchy to limit the number of reasons for testing categories. The hierarchy in the PHS Amsterdam dataset was DCT notification, MCT notification, having symptoms without notification, and unknown. The hierarchy in the first and second RDT studies, was DCT notification, MCT notification, followed by other types of notifications and other types of reasons for testing (*Supplementary methods*). We used the last date of exposure/contact, and the date a test sample was taken, to calculate the exposure-testing interval in all datasets.

**Table 1:**
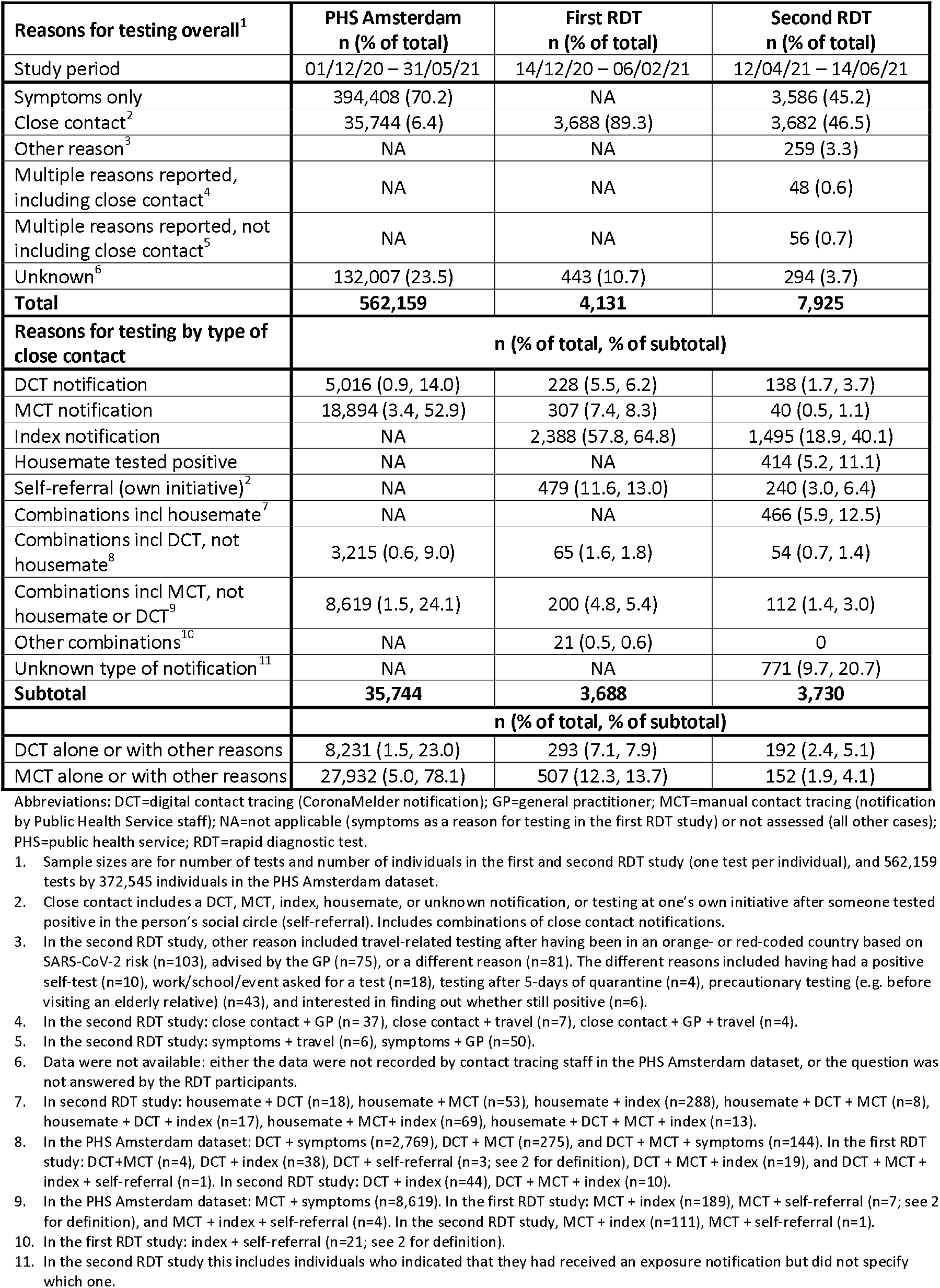
Reasons for testing in the PHS Amsterdam, first RDT, and second RDT studies in the Netherlands, 2020-2021.

| Reasons for testing overall<sup>1</sup> | PHS Amsterdam<br>n (% of total) | First RDT<br>n (% of total) | Second RDT<br>n (% of total) |
| --- | --- | --- | --- |
| Study period | 01/12/20 – 31/05/21 | 14/12/20 – 06/02/21 | 12/04/21 – 14/06/21 |
| Symptoms only | 394,408 (70.2) | NA | 3,586 (45.2) |
| Close contact<sup>2</sup> | 35,744 (6.4) | 3,688 (89.3) | 3,682 (46.5) |
| Other reason<sup>3</sup> | NA | NA | 259 (3.3) |
| Multiple reasons reported, including close contact<sup>4</sup> | NA | NA | 48 (0.6) |
| Multiple reasons reported, not including close contact<sup>5</sup> | NA | NA | 56 (0.7) |
| Unknown<sup>6</sup> | 132,007 (23.5) | 443 (10.7) | 294 (3.7) |
| **Total** | **562,159** | **4,131** | **7,925** |
| **Reasons for testing by type of close contact** | **n (% of total, % of subtotal)** |  |  |
| DCT notification | 5,016 (0.9, 14.0) | 228 (5.5, 6.2) | 138 (1.7, 3.7) |
| MCT notification | 18,894 (3.4, 52.9) | 307 (7.4, 8.3) | 40 (0.5, 1.1) |
| Index notification | NA | 2,388 (57.8, 64.8) | 1,495 (18.9, 40.1) |
| Housemate tested positive | NA | NA | 414 (5.2, 11.1) |
| Self-referral (own initiative)<sup>2</sup> | NA | 479 (11.6, 13.0) | 240 (3.0, 6.4) |
| Combinations incl housemate<sup>7</sup> | NA | NA | 466 (5.9, 12.5) |
| Combinations incl DCT, not housemate<sup>8</sup> | 3,215 (0.6, 9.0) | 65 (1.6, 1.8) | 54 (0.7, 1.4) |
| Combinations incl MCT, not housemate or DCT<sup>9</sup> | 8,619 (1.5, 24.1) | 200 (4.8, 5.4) | 112 (1.4, 3.0) |
| Other combinations<sup>10</sup> | NA | 21 (0.5, 0.6) | 0 |
| Unknown type of notification<sup>11</sup> | NA | NA | 771 (9.7, 20.7) |
| **Subtotal** | **35,744** | **3,688** | **3,730** |
|  | **n (% of total, % of subtotal)** |  |  |
| DCT alone or with other reasons | 8,231 (1.5, 23.0) | 293 (7.1, 7.9) | 192 (2.4, 5.1) |
| MCT alone or with other reasons | 27,932 (5.0, 78.1) | 507 (12.3, 13.7) | 152 (1.9, 4.1) |

Abbreviations: DCT=digital contact tracing (CoronaMelder notification); GP=general practitioner; MCT>manual contact tracing (notification by Public Health Service staff); NA=not applicable (symptoms as a reason for testing in the first RDT study) or not assessed (all other cases); PHS=public health service; RDT=rapid diagnostic test.

Reasons for testing over time were displayed graphically. Population characteristics by reason for testing were tabulated per data source and compared using Pearson’s Chi-squared test for categorical variables and Kruskal-Wallis test for continuous variables. Adjusted standardized residuals of statistically significant categorical variables (p<0.05) were subsequently compared to the critical value, with p-values adjusted by the Bonferroni method. Continuous variables that differed statistically significantly between groups were subsequently analysed by Dunn Kruskal-Wallis multiple comparison test, with p-values adjusted by the Benjamini-Hochberg method. Mean exposure-testing intervals with standard deviations (SD) were calculated by reason for testing and by other population characteristics. Univariable and multivariable Weibull regression and tobit censored regression models were performed to estimate associations between characteristics and exposure-testing intervals. Reason for testing was included in these models as an indicator variable to allow for reporting of multiple reasons per person. Weibull models were a better fit than tobit models because testing peaked on day five after exposure, especially in the first RDT dataset. However, both types of models were fit, with tobit models considered a sensitivity analysis. In the PHS Amsterdam regression models, all individuals for whom an exposure-testing interval could be determined had been part of the MCT programme, and MCT was therefore not included as a variable. All statistical analyses were performed in R versions 3.6.3 or 4.0.3 (R Foundation for Statistical Computing, Vienna, Austria).

## Results

### Reasons for testing

The main reason for testing over time was having symptoms, except in the first RDT study that only included asymptomatic close contacts (Table 1, Figure 2). In the PHS Amsterdam dataset, the overall percentage of test requests due to symptoms was 70.2% over the entire time period, but declined from a daily percentage between 73-97% in the period prior to 1 December 2020 to 60% on 31 May 2021; the overall percentage in the second RDT study that took place in the second quarter of 2021 was 45.2%. Having received a DCT notification, or having been contacted by the MCT programme, were minor reasons for testing in all three datasets over time. The percentages of test requests that listed DCT as one of the reasons was 1.5% in the PHS Amsterdam dataset, 2.4% in the second RDT study, and 7.1% in the first RDT study in asymptomatic close contacts. These percentages were 4.9%, 1.9%, and 12.3% for MCT, respectively. Direct notification by a housemate who tested positive or another index case (any other person who tested positive) was much more common than DCT and MCT notifications: 30% and 58.3% in the second and first RDT studies, respectively. In the first RDT study among asymptomatic close contacts, direct notification by index cases was consistently the main reason for testing over time.

**Figure 2:**
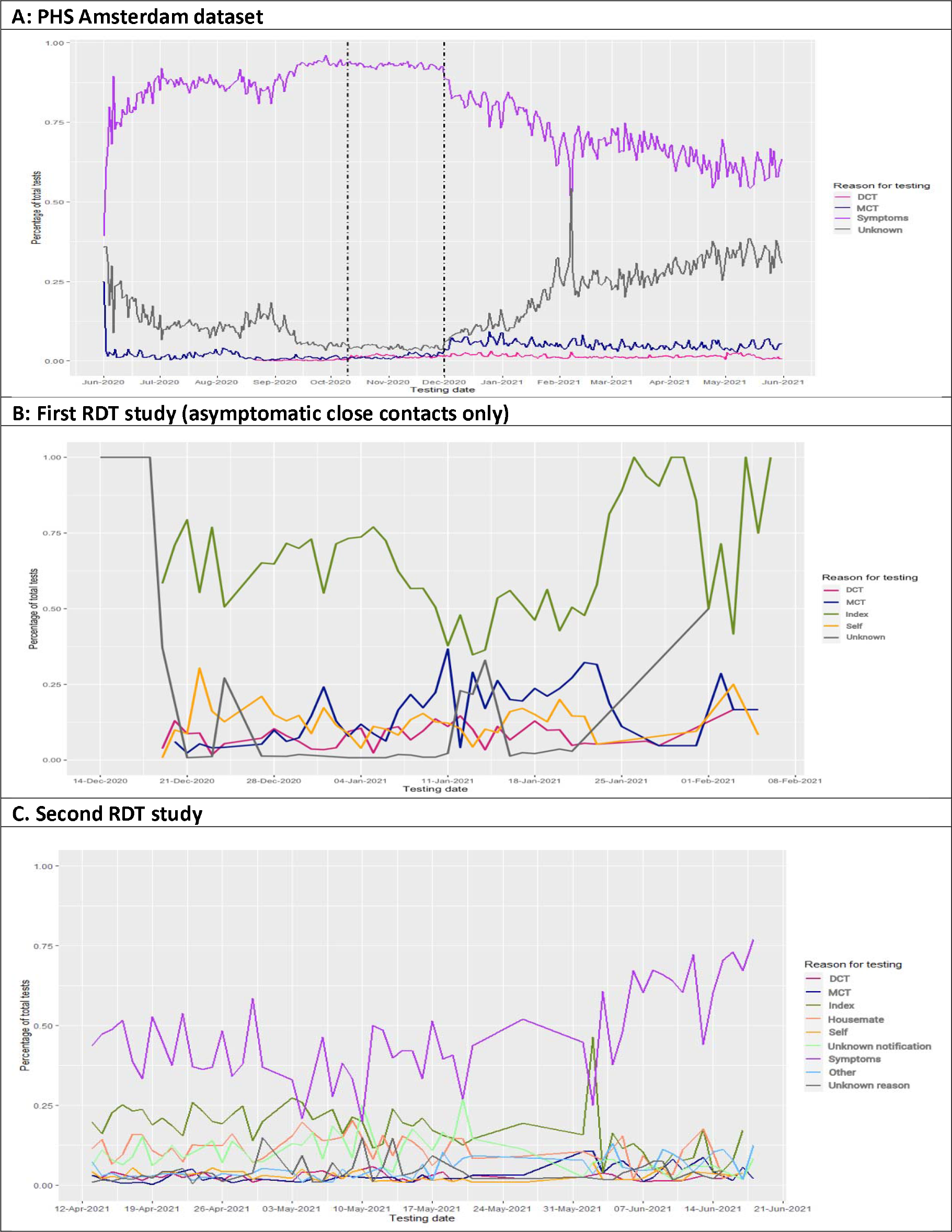
Reasons for testing over calendar time in the PHS Amsterdam, first RDT, and second RDT datasets in the Netherlands, 2020-2021. Abbreviations: DCT=digital contact tracing; Index=a person who tested SARS-CoV-2 positive; MCT=manual contact tracing; PHS=public health service; RDT=rapid diagnostic test; Self=testing at one’s own initiative. In Figure 1A, the first vertical dashed line represents the launch of the CoronaMelder app and the second one the change in testing policy, allowing asymptomatic individuals to get tested after exposure to a close contact. Figure 1A is based on 908,060 tests by 560,775 individuals; 1B on 4,126 tests by 4,126 individuals; and 1C on 7,925 tests by 7,925 individuals.

Most test requests in the PHS Amsterdam dataset (72.2%) and the second RDT study (62.3%) included symptoms as one of the reasons for testing (Tables S1A and S1C). In the first RDT study, 9.2% of asymptomatic close contacts developed symptoms by the time of testing (Table S1B). Furthermore, a substantial proportion of test requesters who reported a DCT or MCT notification also indicated the presence of symptoms (36% or 31% in the PHS Amsterdam dataset and 20% or 28% in the second RDT study, respectively). Test population characteristics (such as age and gender) by reasons for testing are shown in Tables S1A-C and described in the Supplementary results.

### Test positivity percentages

The overall SARS-CoV-2 test positivity was roughly 9% in all datasets (Tables S1A-C). Test positivity in the DCT notification groups was lower than the overall test positivity in the PHS Amsterdam dataset (6.1%), first RDT study (3.4%), and second RDT study (3.7%), but this only reached statistical significance in the former two studies. Test positivity was statistically significantly higher than the overall test positivity in the MCT notification group of the PHS Amsterdam dataset (17.6%), and in the household (22.8%) and unknown notification (13.6%) groups in the second RDT study.

### Mean exposure-testing intervals

In the PHS Amsterdam MCT subset, first RDT study, and second RDT study, the mean exposure-testing intervals were 3.9 (SD 2.6), 4.9 (SD 1.5), and 4.3 (SD 2.0) days, respectively (Tables S2A-C). The DCT notification groups had similar or slightly shorter mean exposure-testing intervals (ranging from 4.2 to 5.2 days) than the MCT notification groups (ranging from 4.5-5.1 days) in all three datasets, but longer intervals than the symptoms group (data available for the PHS Amsterdam MCT subset only; mean 3.1 days) and the household notification group (data available for the second RDT study only; mean 3.1 days). Mean exposure-testing intervals for having been notified by an index case or testing at one’s own initiative were in between those for symptoms or household notification on the one hand and DCT or MCT notification on the other hand (Tables S2A-C). Mean exposure-testing intervals by other test population characteristics are shown in Tables S2A-C and described in the Supplementary results.

### Weibull regression models with exposure-testing interval as outcome

The univariable Weibull models showed that testing after having received a DCT notification was associated with a statistically significantly 4-7% longer exposure-testing interval compared to all other reason for testing groups in all three datasets, but this only remained significant in the multivariable models of the PHS Amsterdam MCT subset and second RDT study (8% and 10%, respectively; Tables 2A-C). In uni- and multivariable Weibull models, testing after MCT notification was associated with a statistically significantly longer interval in the second RDT study (6% and 11%, respectively), testing after index case notification with a shorter interval in the first RDT study (5% and 5%, respectively), testing after a household notification with a shorter interval in the second RDT study (14% and 14%, respectively), and testing at one’s own initiative with a longer interval in the second RDT study (9% and 11%, respectively). In the PHS Amsterdam MCT subset, individuals who were registered as cases or as household contacts had shorter exposure-testing intervals than non-household close contacts with long or short duration of contact (Table 2A). In the multivariable Weibull model excluding cases, testing after a household contact was associated with a significantly shorter interval than testing after a non-household close contacts with long or short duration of contact (Table 2A).

**Table 2A:** Results of Weibull regression models for the interval between last exposure and testing in days – PHS Amsterdam MCT subset.

|  |  | Univariable analysis<sup>1</sup><br>(n<sub>e,t</sub>= 20,647) |  |  | Multivariable analysis<sup>1</sup><br>(n<sub>e,t</sub>= 20,647) |  |  | Multivariable analysis contacts<sup>2</sup><br>(n<sub>e,t</sub>= 15,051) |  |  | Multivariable analysis<br>asymptomatics<sup>3</sup> (n<sub>e,t</sub>= 11,992) |  |  |
| --- | --- | --- | --- | --- | --- | --- | --- | --- | --- | --- | --- | --- | --- |
| Coefficients |  | ETR<sup>4</sup> | 95% CI | p-value | ETR<sup>4</sup> | 95% CI | p-value | ETR<sup>4</sup> | 95% CI | p-value | ETR<sup>4</sup> | 95% CI | p-value |
| **Age:** | 0-14 | 1.10 | 1.08-1.12 | <0.01 | 1.07 | 1.04-1.09 | <0.01 | 1.08 | 1.06-1.10 | <0.01 | 1.06 | 1.04-1.08 | <0.01 |
|  | 15-29 | reference | --- | --- | reference | --- | --- | reference | --- | --- | reference | --- | --- |
|  | 30-44 | 1.02 | 1.00-1.04 | 0.04 | 1.01 | 0.99-1.03 | 0.24 | 1.05 | 1.03-1.07 | <0.01 | 1.03 | 1.01-1.05 | 0.01 |
|  | 45-59 | 1.03 | 1.00-1.05 | 0.01 | 1.03 | 1.01-1.05 | <0.01 | 1.04 | 1.02-1.06 | <0.01 | 1.01 | 0.99-1.04 | 0.24 |
|  | 60+ | 1.08 | 1.05-1.11 | <0.01 | 1.07 | 1.04-1.09 | <0.01 | 1.07 | 1.04-1.10 | <0.01 | 1.07 | 1.04-1.10 | <0.01 |
| **Gender:** | Female | reference | --- | --- | reference | --- | --- | reference | --- | --- | reference | --- | --- |
|  | Male | 1.02 | 1.01-1.04 | <0.01 | 1.02 | 1.00-1.03 | <0.01 | 1.02 | 1.01-1.04 | <0.01 | 1.01 | 1.00-1.03 | 0.07 |
| **Municipality:** | Amsterdam | reference | --- | --- | reference | --- | --- | reference | --- | --- | reference | --- | --- |
|  | Surrounding | 0.99 | 0.97-1.00 | 0.12 | 0.98 | 0.96-0.99 | <0.01 | 0.97 | 0.96-0.99 | <0.01 | 0.99 | 0.97-1.00 | 0.21 |
| **Type of contact:<sup>5</sup>** |  |  |  |  |  |  |  |  |  |  |  |  |  |
| Household |  | reference | --- | --- | reference | --- | --- | reference | --- | --- | reference | --- | --- |
| Close, long |  | 1.07 | 1.05-1.09 | <0.01 | 1.07 | 1.06-1.09 | <0.01 | 1.06 | 1.04-1.08 | <0.01 | 1.01 | 1.00-1.03 | 0.10 |
| Close, short |  | 1.09 | 1.02-1.17 | <0.01 | 1.10 | 1.03-1.17 | <0.01 | 1.08 | 1.02-1.14 | 0.01 | 1.06 | 0.99-1.13 | 0.09 |
| Other contact |  | 1.00 | 0.86-1.17 | 0.99 | 1.00 | 0.85-1.16 | 0.98 | 0.98 | 0.85-1.13 | 0.77 | 1.01 | 0.86-1.20 | 0.88 |
| Case |  | 0.87 | 0.86-0.89 | <0.01 | 0.91 | 0.89-0.92 | <0.01 | Not incl | --- | --- | 0.88 | 0.87-0.90 | <0.01 |
| **DCT:** | No | reference | --- | --- | reference | --- | --- | reference | --- | --- | reference | --- | --- |
|  | Yes | 1.06 | 1.00-1.12 | 0.05 | 1.08 | 1.01-1.14 | 0.02 | 1.09 | 1.02-1.16 | <0.01 | 1.10 | 1.03-1.18 | <0.01 |
| **Symptoms:** | No | reference | --- | --- | reference | --- | --- | reference | --- | --- | Not | --- | --- |
|  | Yes | 0.84 | 0.83-0.85 | <0.01 | 0.86 | 0.85-0.88 | <0.01 | 0.87 | 0.85-0.88 | <0.01 | included | --- | --- |
| **Test result:** | Negative | reference | --- | --- | Not | --- | --- | Not | --- | --- | Not | --- | --- |
|  | Positive | 0.89 | 0.88-0.91 | <0.01 | included<sup>6</sup> | --- | --- | included<sup>6</sup> | --- | --- | included<sup>6</sup> | --- | --- |

Abbreviations: CI=confidence interval; DCT=digital contact tracing; ETR=event-time ratio; Index=a person who tested SARS-CoV-2 positive; MCT>manual contact tracing; PHS=public health service; RDT=rapid diagnostic test; Self=testing at one's own initiative.
1. Based on 20,647 exposure- testing intervals (n<sub>e,t</sub>) by 20,355 individuals (n<sub>i</sub>) between 1 December 2020- 31 March 2021. Missing values for type of contact (n<sub>e,t</sub>= 69), test result (n<sub>e,t</sub>= 78), and gender (n<sub>e,t</sub>= 47). 2. Based on 15,051 exposure-test intervals (n<sub>e,t</sub>) in 14,827 individuals (n<sub>i</sub>). Missing values for type of contact (n<sub>e,t</sub>= 69), test result (n<sub>e,t</sub>= 60) and gender (n<sub>e,t</sub>= 27). 3. Based on 11,992 exposure-test intervals (n<sub>e,t</sub>) in 11,855 asymptomatic individuals (n<sub>i</sub>). Missing values for type of contact (n<sub>e,t</sub>= 40), test result (n<sub>e,t</sub>= 47) and gender (n<sub>e,t</sub>= 28). 4. The outcome measure is event time ratio (ETR). An ETR of 1.08 for having received a DCT notification can be interpreted as the relative increase in the time interval between exposure and testing by approximately 8 percent as compared to not having received a DCT notification. 5. "Close" is defined as within 1.5 meters of an infectious person; "long" as more than 15 minutes; "short" as 15 minutes or less but with high intensity (e.g. coughing in someone's face, kissing); "household" as a close contact within the same residence; and "other" as any other contact with an infectious person. 6. Test result was highly correlated with contact type and was therefore not included in the multivariable models.

**Table 2B:** Results of Weibull regression models for the interval between last exposure and testing in days – first RDT study.

|  |  | Univariable analysis<sup>1</sup> (n= 3,646) |  |  | Multivariable analysis<sup>1</sup> (n= 3,646) |  |  |
| --- | --- | --- | --- | --- | --- | --- | --- |
| Coefficients |  | ETR<sup>2</sup> | 95% CI | p-value | ETR<sup>2</sup> | 95% CI | p-value |
| **Age:** | 16-29 | reference | --- | --- | reference | --- | --- |
|  | 30-44 | 1.02 | 1.00-1.04 | 0.13 | 1.02 | 1.00-1.04 | 0.11 |
|  | 45-59 | 1.00 | 0.98-1.02 | 0.95 | 0.99 | 0.97-1.01 | 0.58 |
|  | 60+ | 1.03 | 1.01-1.06 | <0.01 | 1.02 | 1.00-1.05 | 0.04 |
| **Gender:** | Female | reference | --- | --- | reference | --- | --- |
|  | Male | 1.00 | 0.98-1.02 | 0.97 | 1.00 | 0.98-1.01 | 0.92 |
| **Testing region:** | Brabant | reference | --- | --- | reference | --- | --- |
|  | Rotterdam | 0.96 | 0.94-0.97 | <0.01 | 0.97 | 0.95-0.98 | <0.01 |
| **DCT:** | No | reference | --- | --- | reference | --- | --- |
|  | Yes | 1.04 | 1.01-1.07 | <0.01 | 1.00 | 0.96-1.04 | 0.96 |
| **MCT:** | No | reference | --- | --- | reference | --- | --- |
|  | Yes | 1.02 | 1.00-1.05 | 0.05 | 1.00 | 0.97-1.02 | 0.76 |
| **Index:** | No | reference | --- | --- | reference | --- | --- |
|  | Yes | 0.95 | 0.94-0.97 | <0.01 | 0.95 | 0.92-0.98 | <0.01 |
| **Self:** | No | reference | --- | --- | reference | --- | --- |
|  | Yes | 1.02 | 1.00-1.04 | 0.12 | 0.97 | 0.94-1.01 | 0.16 |
| **Unknown Contact:** | No | reference | --- | --- | reference | --- | --- |
|  | Yes | 1.00 | 0.92-1.11 | 0.86 | 0.97 | 0.87-1.07 | 0.51 |
| **Symptoms:** | No | reference | --- | --- | reference | --- | --- |
|  | Yes | 1.02 | 0.99-1.05 | 0.14 | 1.03 | 1.00-1.06 | 0.03 |
| **Test result:** | Negative | reference | --- | --- | reference | --- | --- |
|  | Positive | 0.98 | 0.96-1.01 | 0.25 | 0.98 | 0.96-1.01 | 0.25 |

Abbreviations: CI=confidence interval; DCT=digital contact tracing; ETR=event-time ratio; Index=a person who tested SARS-CoV-2 positive; MCT>manual contact tracing; PHS=public health service; RDT=rapid diagnostic test; Self=testing at one's own initiative.
1. Includes 3,646 exposure-test intervals in 3,646 participants between 14 December 2020 - 6 February 2021. Only participants who reported a close contact were asked the date of last exposure and dates are missing (n=480) or not logical (before testing date or more than 14 days after testing, n=5). Additional missing values for symptoms (n=21), age (n=9), gender (n=14), and test location (n=9). 2. The outcome measure is event time ratio (ETR). An ETR of 1.04 for having received a DCT notification can be interpreted as the relative increase in the time interval between exposure and testing by approximately 4 percent as compared to not having received a DCT notification.

**Table 2C:** Results of Weibull regression models for the interval between last exposure and testing in days – second RDT study.

|  |  | Univariable analysis<sup>1</sup> (n=3,172) |  |  | Multivariable analysis<sup>1</sup> (n=3,172) |  |  | Multivariable among asymptomatic<sup>2</sup> (n=2,179) |  |  |
| --- | --- | --- | --- | --- | --- | --- | --- | --- | --- | --- |
| Coefficients |  | ETR<sup>3</sup> | 95% CI | p-value | ETR<sup>3</sup> | 95% CI | p-value | ETR<sup>3</sup> | 95% CI | p-value |
| **Age:** | 16-29 | reference | --- | --- | reference | --- | --- | reference | --- | --- |
|  | 30-44 | 1.03 | 1.00-1.06 | 0.08 | 1.01 | 0.98- 1.04 | 0.69 | 1.00 | 0.96- 1.03 | 0.92 |
|  | 45-59 | 1.01 | 0.97- 1.04 | 0.73 | 1.01 | 0.97- 1.04 | 0.70 | 0.99 | 0.96- 1.03 | 0.76 |
|  | 60+ | 1.03 | 0.98-1.08 | 0.20 | 0.98 | 0.93- 1.02 | 0.32 | 0.95 | 0.90- 1.00 | 0.07 |
| **Gender:** | Female | reference | --- | --- | reference | --- | --- | reference | --- | --- |
|  | Male | 0.99 | 0.96- 1.01 | 0.35 | 0.99 | 0.97-1.02 | 0.49 | 0.99 | 0.96- 1.02 | 0.38 |
| **Testing region:** | Brabant | reference | --- | --- | reference | --- | --- | reference | --- | --- |
|  | Rotterdam | 0.99 | 0.96-1.02 | 0.62 | 0.99 | 0.96- 1.02 | 0.68 | 0.98 | 0.95- 1.01 | 0.23 |
|  | Zwolle | 0.96 | 0.93- 1.00 | 0.03 | 0.95 | 0.91-0.98 | <0.01 | 0.97 | 0.93- 1.00 | 0.07 |
| **DCT:** | No | reference | --- | --- | reference | --- | --- | reference | --- | --- |
|  | Yes | 1.07 | 1.02-1.13 | <0.01 | 1.10 | 1.04 - 1.16 | <0.01 | 1.05 | 0.99- 1.11 | 0.08 |
| **MCT:** | No | reference | --- | --- | reference | --- | --- | reference | --- | --- |
|  | Yes | 1.06 | 1.01- 1.10 | 0.02 | 1.11 | 1.06 -1.16 | <0.01 | 1.08 | 1.03- 1.13 | <0.01 |
| **Index:** | No | reference | --- | --- | reference | --- | --- | reference | --- | --- |
|  | Yes | 1.03 | 1.00- 1.05 | 0.04 | 1.04 | 0.99- 1.08 | 0.10 | 1.02 | 0.98- 1.07 | 0.30 |
| **Housemate:** | No | reference | --- | --- | reference | --- | --- | reference | --- | --- |
|  | Yes | 0.86 | 0.84- 0.89 | <0.01 | 0.86 | 0.83- 0.90 | <0.01 | 0.92 | 0.88- 0.95 | <0.01 |
| **Self:** | No | reference | --- | --- | reference | --- | --- | reference | --- | --- |
|  | Yes | 1.09 | 1.03-1.16 | <0.01 | 1.11 | 1.03 -1.19 | <0.01 | 1.12 | 1.03- 1.21 | <0.01 |
| **Unknown Contact:** | No | reference | --- | --- | reference | --- | --- | reference | --- | --- |
|  | Yes | 1.02 | 0.98- 1.05 | 0.29 | 1.03 | 0.97- 1.08 | 0.34 | 1.05 | 0.99- 1.11 | 0.08 |
| **Symptoms:** | No | reference | --- | --- | reference | --- | --- | Not included | --- | --- |
|  | Yes | 0.98 | 0.95- 1.00 | 0.08 | 0.98 | 0.95- 1.01 | 0.22 |  |  |  |
| **Test result:** | Negative | reference | --- | --- | reference | --- | --- | reference | --- | --- |
|  | Positive | 0.93 | 0.89- 0.96 | <0.01 | 0.95 | 0.92- 0.99 | 0.02 | 0.92 | 0.87- 0.97 | <0.01 |
| **Vaccinated:** | No | reference | --- | --- | reference | --- | --- | reference | --- | --- |
|  | Yes | 1.04 | 1.00- 1.08 | 0.07 | 1.03 | 0.99- 1.08 | 0.16 | 1.04 | 0.99- 1.09 | 0.09 |
| **Previous infection:** | No | reference | --- | --- | reference | --- | --- | reference | --- | --- |
|  | Yes | 1.00 | 0.96- 1.04 | 0.95 | 1.00 | 0.95- 1.04 | 0.84 | 0.98 | 0.94- 1.03 | 0.37 |

Abbreviations: CI=confidence interval; DCT=digital contact tracing; ETR=event-time ratio; Index=a person who tested SARS-CoV-2 positive; MCT>manual contact tracing; PHS=public health service; RDT=rapid diagnostic test; Self=testing at one's own initiative.
1. Includes 3,172 exposure-test intervals in 3,172 participants between 12 April- 14 June 2021. Only participants who reported a close contact were asked the date of last exposure and dates are missing (n=519) or not logical (before testing date or more than 14 days after testing, n=39). Additional missing values for symptoms (n=13), age (n= 6), gender (n=9), vaccination status (n=1), and prior SARS-CoV-2 infection (n= 9). 2. 2,179 exposure-test intervals in 2,179 participants. Missing values for age (n= 4), gender (n=7), and prior SARS-CoV-2 infection (n= 3). 3. The outcome measure is event time ratio (ETR). An ETR of 1.04 for having received a DCT notification can be interpreted as the relative increase in the time interval between exposure and testing by approximately 4 percent as compared to not having received a DCT notification.

In the PHS Amsterdam MCT subset, testing because of symptoms was associated with statistically significantly shorter exposure-testing intervals (13-16%) in all regression models (Table 2A). In the second RDT study, intervals were not available for individuals who tested because of having symptoms. In the group who tested because of having been exposed to a positive individual, reporting symptoms was not significantly associated with a shorter interval in the univariable and/or multivariable models but trends in that direction were apparent (Table 2C).

In the PHS Amsterdam MCT subset, individuals who ended up testing positive had a 11% shorter exposure-test interval than those who ended up testing negative (Table 2A). Similar trends were seen in the first and second RDT study (Tables 2B-C), but only reaching significance in the second RDT study (7% shorter in univariable and 5% shorter in the multivariable models). In the second RDT study, COVID-19 vaccination status and previous SARS-CoV-2 infection were not significantly associated with exposure-testing intervals (Table 2C). The tobit censored regression model results are shown in Tables S3A-C and described in the Supplementary results.

## Discussion

We found that the epidemiological impact of both DCT and MCT were limited during the Dutch 2020-2022 SARS-CoV-2 epidemic. Only around 2% of all tests took place after a DCT notification, and 2-5% after a MCT notification depending on MCT capacity at the time. Additionally, test positivity among those testing after a DCT notification was significantly lower than among individuals testing for other reasons, the exposure-test intervals were longer after a DCT or MCT notification than for other reasons for testing, and about 20-36% of those who had received a DCT or MCT notification had symptoms at the time of test request and might have tested anyway even without having received the notification. These findings are in line with a self-evaluation performed by the Dutch PHS (GGD-GHOR) that found that the majority of tests at PHS test sites were not triggered by MCT or DCT notification, and test positivity was lower than average after DCT notification (13).

By the end of May 2021, almost 30% of the Dutch population had downloaded the CoronaMelder app but only around 18% were still actively using it (5). These percentages were similar to those in other countries that rolled out voluntary DCT: DCT apps were downloaded by 28% of the Norwegian population (14), 28% of the UK population (15), 26% of the Swiss population (declining to 22% active users over time) (16), 17% of the Canadian population (17), and use was reported by 21% of cases in Finland and 22% of cases in New South Wales, Australia (18). Direct comparisons between epidemiological impact study results in these countries are not possible because of differences in how the contact-tracing apps were embedded within existing public health infrastructures, availability of data, and the degree of empirical evidence versus mathematical modelling used to estimate impact. However, overall, they showed only modest (14,15,19) to limited impact (5,18,20– 22), with results being least pronounced when conclusions were predominantly based on empirical evidence.

Most impact studies cited low uptake as the main reason for the limited DCT impact, and several barriers to adoption have been identified: concerns about cybersecurity and privacy; lower levels of education, income, expectation about efficacy/perceived benefits, self-efficacy, and trust in government; higher age; and living in a community with lower levels of use (5,23–28). Several of these barriers are amenable to intervention, such as education and promotion campaigns. These campaigns were suboptimal in the Netherlands (5). In addition, combining DCT apps with other practical functionalities might have improved uptake. For example, the UK National Health Service (NHS) COVID-19 app included multiple functionalities and fared better than CoronaMelder: it was still regularly being used by around 28% of the British population by the end of December 2021 (15). These functionalities included a symptom checker connected to test requesting, a QR-scanner for checking into certain venues, and current government SARS-CoV-2 prevention guidelines in the local area. The Dutch government considered combining the functionalities of the CoronaMelder and CoronaCheck apps (to generate vaccination, test, and/or cure certificates) into one app, as has been done by other European governments such as Germany (Corona-Warn-App), Austria (The Stopp Corona App), and France (TousAntiCovid) (20,29), but eventually decided against this. This may have been a missed opportunity. The CoronaCheck app had been downloaded by 90% of the Dutch population by 1 March 2022 (30).

Our analyses suggest that the PHS MCT programme did not contribute much to epidemic control either. Unfortunately, we could not compare these findings to the above-mentioned impact studies because those studies did not place DCT in the wider context of other reasons for testing. In our study, most people in all three datasets tested because of having symptoms or because they themselves found out that someone in their social circle had tested positive. While MCT will likely continue to play an important role in the control of infectious diseases, its utility in the context of respiratory viruses that rapidly transmit within general populations should be re-evaluated. For example, instead of trying to maintain MCT at population level after containment has failed (which was regularly the case in the 2020-2022 Dutch SARS-CoV-2 epidemic), more focussed uses might be more appropriate and cheaper.

Unexpectedly, we found that DCT and MCT did not shorten delays between last exposure and testing compared to symptom-based testing. We hypothesise that suboptimal testing policies and CoronaMelder procedures are partly to blame for this. Between 1 December 2020 and 18 February 2022, exposed but asymptomatic individuals were advised to test five days after exposure, hence the fairly uniform distribution of 5-day intervals in the first RDT study. Furthermore, when a CoronaMelder user tested positive, they had to wait for a PHS employee to contact them to be able to notify their contacts. A Swiss study reported that non-household MCT contacts who had also received a SwissCovid app notification generally went into quarantine a day earlier than non-household MCT contacts who had not received one (31). Nevertheless, the Swiss team also identified unnecessary delays due to several bottlenecks in the exposure-notification-action cascade, such as delayed delivery of codes for test result sharing, complex user interfaces, and misaligned incentives for subsequent mitigation behaviours (16).

The main limitations of our study relate to data availability and quality. To safeguard privacy, the only publicly accessible CoronaMelder-generated data are anonymous data on app downloads and active use (the number of smartphones connecting with the backend server) (5). We therefore had to use proxy data sources. First, all three datasets that we used consisted mostly of self-reported data, which can suffer from misreporting and/or recall bias. For example, the last exposure date was not always available. Individuals who booked a test online had to consistently answer questions about CoronaMelder use to be able to make a booking, but these questions were less consistently asked and recorded when individuals booked a test by phone or within the MCT process (32). Second, test policies evolved over time. We already mentioned the day-5 test policy for individuals without symptoms. In addition, from 31 March 2021 onwards, self-testing became increasingly available and popular. The effects of this on our analyses are, however, limited because the three studies included in this report ran until mid-June 2021, before self-testing really took off. Third, we relied heavily on HPZone data for some of the PHS Amsterdam analyses, which limited the study population to individuals who had participated in the MCT programme. Furthermore, MCT capacity and guidelines fluctuated over time. Finally, we attempted to control for confounding in the multivariable models, but residual confounding cannot be excluded.

We conclude that both DCT and MCT had limited impacts on the Dutch 2020-2022 SARS-CoV-2 epidemic. However, in future epidemics, the impact of DCT might be improved by concerted efforts to increase app use coverage, elimination of contact-tracers from the digital notification process to minimise delays, and ensuring that everyone who is notified can get tested right away, including by self-testing.

## Data Availability

PHS Amsterdam data are not publicly available for privacy reasons. Individual participant data of the two RDT studies will be made available, after deidentification, to researchers who provide a methodologically sound proposal. Proposals should be directed to Professor Carl Moons. Data requestors will need to sign a data access agreement.

## Acknowledgments

We thank our colleagues at the various Public Health Services and associated laboratories, the Julius Center for Health Sciences and Primary Care at the University Medical Center Utrecht, the National Institute for Public Health and the Environment (RIVM), and the Erasmus University Medical Center who contributed to the studies described in this paper.

## Author contributions

JHHMvdW, AM, and JB initiated and designed the study. WTH assembled and analysed the data with input from JB, AM, and JHHMvdW. ES, RV, SvdH, JK, CM, and JHHMvdW designed the questionnaires of the first and second RDT studies and implemented those studies. WTH and JHHMvdW wrote the first draft of this manuscript and all other authors critically read the manuscript and provided feedback.

## Supporting information captions

Supplementary introduction

Supplementary methods

Supplementary results

Figure S1: CoronaMelder smartphone application

Figure S2: Participant flow diagram PHS Amsterdam datasets

Figure S3: Questionnaires used in the first RDT and second RDT studies (in Dutch language)

Figure S4: Participant flow diagrams first and second RDT studies

Table S1A: Test population characteristics by reason for testing – PHS Amsterdam

Table S1B: Test population characteristics by reason for testing – first RDT study (asymptomatic close contacts)

Table S1C: Test population characteristics by reason for testing – second RDT study

Table S2A: Mean (SD) interval in days between first exposure and testing – PHS Amsterdam MCT subset

Table S2B: Mean (SD) interval in days between last exposure and testing – first RDT study (asymptomatic close contacts)

Table S2C: Mean (SD) interval in days between last exposure and testing – second RDT study

Table S3A: Tobit regression model for exposure-test intervals – PHS Amsterdam MCT subset

Table S3B: Tobit regression model for the exposure-test intervals – first RDT study

Table S3C: Tobit regression model for the exposure-test intervals – second RDT study

