## Supplement for "The epidemiological impact of digital and manual contact tracing on the SARS-CoV-2 epidemic in the Netherlands: empirical evidence"

**Supplementary INTRODUCTION**

*Milestones in the Dutch SARS-CoV-2 epidemic during the data collection period*

The data collection period was from 1 June 2020 to 14 June 2021 and some important milestones related to SARS-CoV-2 governmental measures, testing policies, and manual contact tracing (MCT) phases took place in the Netherlands during this time period.^1^ The original Wuhan virus variant circulated from February to December 2020: the first confirmed case was announced on 27 February and the first death on 6 March.^2^ Travel and contact reduction measures were gradually introduced between 9 March and 28 April 2020 and gradually relaxed thereafter, with the peak of this first wave occurring on 8 April 2020. Public testing was introduced on 1 June 2020 (see below). A second wave occurred between late September 2020 and late January 2021. During this wave, the Wuhan virus variant was gradually replaced by the Alpha variant, which remained in circulation until mid-August 2021. A partial lockdown came into effect on 14 October 2020 and a strict lockdown (with a maximum of two visitors allowed per day, all non-essential shops closed, and all education online) on 15 December 2020. The Dutch COVID-19 vaccination campaign started on 6 January 2021. On 23 January 2021, a curfew was imposed from 9 pm until 4:30 am. Between 1 March and 5 June 2021, restrictions were gradually lifted as vaccination levels increased. After the data collection period, short epidemic waves due to the Delta variant occurred in July 2021 and October/November 2021, and a massive wave due to Delta and Omicron variants from December 2021 until April 2022.

SARS-CoV-2 testing at Public Health Service (PHS) sites became available for anyone with symptoms from 1 June 2020 onwards.^1,3,4^ Close contacts without symptoms were advised to get tested on the fifth day after their last contact with an index case between 1 December 2020 and 18 February 2021, and to get tested as soon as possible after exposure notification as well as on the fifth day since last contact from 18 February 2021 onwards. Commercial test sites catering to asymptomatic individuals that needed proof of a negative test in order to gain access to events or for travel were available from February 2021 until 23 March 2022. On 31 March 2021, the first SARS-CoV-2 rapid antigen self-tests became available in pharmacies and stores.^5^ Until February 2022, individuals with a positive self-test were asked to do a repeat test at a PHS test site. Testing at PHS sites was almost exclusively by PCR test throughout the epidemic. Commercial testing could be done by either PCR or antigen lateral flow test, depending on the access or travel requirements of the person requesting a test. Self-testing was exclusively by antigen lateral flow test. Results of PCR tests initially took a few days to become available but were available within two days later on in the epidemic. The sensitivity and specificity of these tests was very high for all virus variants. Later flow tests produced a result immediately but were less sensitive and specific, especially in asymptomatic individuals.^6,7^

Implementation of the MCT programme was phased regionally depending on regional infection pressure and MCT capacity.^4^ These phases were numbered one to five. Phase 1 consisted of professional notification of all contacts reported by each index case and complete surveillance data collection. We considered MCT phases 3-5 ‘scaled-down’ because in these phases, index cases were asked to notify their contacts themselves and only limited surveillance data was collected. In the period covering the PHS Amsterdam dataset (1 December 2020 - 31 May 2021), the MCT programme in Amsterdam was scaled down between 7 December 2020 and 13 January 2021 and again between 15 March and 19 May 2021. During the first RDT study (14 December 2020 - 6 February 2021), the MCT programme in Rotterdam was scaled down until 30 December 2020 and in West-Brabant until 6 January 2021. During the second RDT study (12 April - 18 June 2021), the MCT programmes in West-Brabant and Rotterdam (but not Zwolle) were scaled down between 12 April and 19 May 2021.

#### *CoronaMelder smartphone application technical details*

The Dutch government passed the Temporary Act Notification Application COVID-19 (Tijdelijke Wet Notificatieapplicatie Covid-19 in Dutch) on 6 October 2020, and launched the national contact tracing app dubbed CoronaMelder on 10 October 2020.^8^ The app used the decentralized protocol of the Google/Apple Exposure Notification (GAEN) Application Programming Interface (API).^9^ GAEN was inspired by the Decentralized Privacy-Preserving Proximity Tracing (DP-3T) protocol, which was developed by an international group of technologists, engineers, epidemiologists, and legal experts.^10,11^ CoronaMelder was developed by a team commissioned by the Dutch Ministry of Health, Welfare and Sports in the summer of 2020.

The app generated a 16-byte random string called Temporary Exposure Key (TEK) approximately every 24 hours. TEKs were transformed into Rolling Proximity Identifiers (RPIs) using the hash function and the AES algorithm, and these RPIs were broadcast through Bluetooth Low Energy by the user’s smartphone. The RPIs changed every time the Bluetooth Low Energy randomised address changed (every 15 minutes on average). In this way, both TEKs and RPIs could not be traced back to individuals.^10^ RPIs of other CoronaMelder users, together with Received Signal Strength Indicators (a proxy for the distance between two smartphones) and the cumulative contact duration, were stored locally on the user’s phone for 14 days. Additionally, the user’s own TEKs were stored on the phone for 14 days. This duration of 14 days was based on the assumption that SARS-CoV-2 could potentially be transmitted over a period of 14 days after exposure. When smartphones with CoronaMelder in active mode came into contact, RPIs together with the indicators of distance and duration of contact were exchanged.

When MCT capacity was optimal (phase 1), PHS staff phoned all individuals who had tested positive at a test site and asked them if they were using CoronaMelder. If yes, they asked the user if they would like to share their unique PHS-key as shown in the app (Figure S1). If yes, the PHS employee added the date of symptoms onset (or of the positive test, if no symptoms were present) to this key. The app user subsequently received a notification (a blue pop-up button), which s/he could (voluntarily) press to upload their TEKs to the backend server. From 11 November 2021 onwards (which is after the three datasets used in this study had already been collected), the app user could upload their PHS-key themselves rather than having to contact the PHS service first.^12^ The backend server was managed by KPN, a Dutch telecommunications company, via CIBG, which is an implementing organization of the Dutch Ministry of Health, Welfare and Sports. After uploading, the TEKs were converted to Diagnostic Keys (DKs) and stored on the backend server for 14 days. Each time an app user’s smartphone connected to the server (approximately six times per day), the app downloaded the list of current DKs and their corresponding RPIs and compared them to the RPIs that were saved on the phone. If there was a match, the proximity, contact duration, and infectious window (based on symptom onset or positive test date) of the infected close contact were used to determine whether transmission may have occurred. If this was the case, the app user received an exposure notification (Figure S1), showing the date of this potential transmission and advice on what to do next. Once the app user had read the notification, it was automatically removed from their phone permanently.

*The exposure notification cascade*

For DCT (and MCT) to have epidemiological impact, all steps of the exposure notification cascade should be optimised. Figure 1 in the main manuscript visualises this cascade. It consists of two separate yet interconnected cascades: part A and part B. An individual enters the cascade after downloading the app (A1). Some individuals may drop out of the cascade after this step because they uninstall the app or have their Bluetooth Low Energy (always or intermittently) turned off. In A2, CoronaMelder is active and exchanging RPIs with other app users through Bluetooth Low Energy. App users spent most of their time in A1 and A2 with the app running in the background. At some point, an exposure notification could have been triggered by CoronaMelder (A3), but also by the MCT programme (B2), by inner circle notification (hearing via the inner circle that an exposure might have happened, either at the request of PHS staff when MCT was scaled down (B3) or spontaneously (A11)), or by self-referral (e.g. having COVID-19-like symptoms; A12). After a trigger, the general guidance throughout the epidemic (also included in CoronaMelder notifications) was to immediately go into quarantine (A5-6), to get tested (A7-9), and/or to request medical help if needed (A10). Exposed individuals were advised to stay in quarantine until receiving a negative test result or for a minimum of 10 days after the exposure (A6). They were advised to get tested as soon as possible after symptom onset; testing without symptoms became possible after 1 December 2020 (A7; see also above).

Individuals testing SARS-CoV-2-positive entered part B of the cascade. The app user who tested positive contacted, or was contacted by, a PHS employee (B1), who either conducted MCT (B2-3; depending on the MCT capacity at the time) and/or facilitated DCT (B3-4; this remained possible even if the MCT was scaled down). When the MCT programme was functioning optimally (phase 1), each person who tested positive was phoned by a PHS employee (B1) and this PHS employee also phoned the index cases’ identifiable close contacts (B2). However, when the MCT programme was scaled down, index cases were asked to warn their identifiable close contacts themselves (B3). Unidentifiable close contacts could only be warned via the DCT (B5) as described above.

**SUPPLEMENTARY METHODS**

*Description of data sources*

We could not use data collected via the CoronaMelder application itself due to the privacy sensitive configuration of the app. Instead, we used routinely collected public health data at the PHS Amsterdam and data from two SARS-CoV-2 rapid diagnostic test (RDT) accuracy studies that were conducted at various other PHS test sites in the Netherlands.^6,7^

PHS Amsterdam data was extracted from the national CoronIT, HPZone, and Osiris databases. CoronIT is a national database established by the Dutch Ministry of Health, Welfare, and Sports specifically for use during the SARS-CoV-2 pandemic. The CoronIT database includes test request data such as age, gender, postal code, reason for testing, having symptoms (and if yes, which ones) at the time of test request, the date/time of test appointment, the date/time of testing, and the test result. HPZone is the MCT database that is used by PHS MCT programmes throughout the country. The PHS Amsterdam HPZone data that we had access to for this study included whether an individual was a case or a contact (the status of contacts who subsequently tested positive changed from contact to case), exposure date, test result, and sociodemographic information. Osiris is the national surveillance database of the National Institute of Public Health and the Environment where all notifiable infections (including SARS-CoV-2) are reported. The only Osiris variable that we used was the level of exposure of those contacted via the MCT program (household contact, close contact of long duration, close contact of short duration, and other contact). At the PHS Amsterdam, for each individual who tested positive, HPZone and Osiris data were entered at the same time by the same employee.

The PHS Amsterdam dataset containing only CoronIT data between 1 December 2020 and 31 May 2021 included 562,159 tests (nt) by 372,545 individuals (ni) (Figure S2). For the exposure-testing interval analyses, CoronIT data were merged with HPZone and Osiris data because exposure dates were only available in HPZone and levels of exposure only in Osiris. This subset of the PHS Amsterdam dataset (referred to as the PHS MCT subset because all individuals in this dataset had been part of the MCT programme as either a case or a contact or both) included 20,647 exposure-testing intervals (n_e-t_) by 20,355 individuals (n_i_) (Figure S2). The merging process was as follows. HPZone data was selected based on whether an exposure date (ne) was present. A subset was made for the period in which the exposure took place two weeks before 1 December 2020 (17 November 2020) and before 31 March 2021 due to data availability. Duplicate entries were removed. Next, the Osiris variable for level of exposure was added using personal identifiers for merging with HPZone. Duplicate entries based on personal identifiers and degree of contact were removed. The CoronIT dataset between 1 December 2020 and 31 March 2021 was used to add the test date (t), again using personal identifiers for merging. Some individuals had multiple exposure- testing intervals (ne-t). For example, one individual was registered as a contact in HPZone at two different time points and got tested a total of 6 times across the duration of the study period. For this person, 12 exposure-testing intervals were present in the combined dataset. After merging, the combined dataset was cleaned. Entries in which the exposure date was after the testing date were removed. When multiple test-dates and one exposure date were present, the most relevant test-date was selected and the others removed. For example, when two test dates were far apart, only the test date closest to the exposure date was kept. When multiple tests were done soon after one exposure, only the first test-date after the exposure was kept. Furthermore, when exposure-testing intervals were more than 14 days, the test-date was set to missing. We assumed that in these cases, another reason than the exposure recorded in the database likely triggered the testing. Finally, entries with an unknown exposure-testing interval were removed from the dataset because the reason for testing was not available for these cases.

The RDT studies were previously published and the logistics summarised in the main manuscript.^6,7^ We used data from the RDT study-specific questionnaires (Figure S3). The first RDT study dataset contained 4,126 individuals (Figure S4). The second RDT study dataset contained 7,925 individuals but date of last exposure, and thereby an exposure-testing interval, was only available for 3,172 individuals (Figure S4). The participants had already completed the routine PHS questionnaire when booking a test online, or answered questions by PHS staff over the phone, but that routine data was not available to us for this study. The data from the two or three regions participating in each RDT study, respectively, were combined to form one dataset per study. In the first RDT study, the reason for testing question had four answer options (tick all that apply): notified via CoronaMelder, MCT, or index case, or testing at one’s own initiative because someone in the social circle tested positive (coded as ‘self’); the study included only individuals who were asymptomatic at test request and therefore did not ask about symptoms as a reason for testing. In the second RDT study, the reason for testing question had five main answer options (tick all that apply): having symptoms, having been in contact with someone who tested positive, the individual’s general practitioner (GP) advised testing, having travelled to an orange/red country, or other. Individuals who ticked the second option were additionally asked for a last date of contact with the index case, and for the following types of notification/contacts: CoronaMelder, MCT, index case, housemate, or testing at one’s own initiative (‘self’). The ‘self’ category in the first RDT study likely includes both the housemate and ‘self’ categories of the second RDT study.

*Supplementary statistical analyses*

In all datasets, individuals could report multiple reasons for testing, and we used a categorisation hierarchy to limit the number of reasons for testing categories. The hierarchy in the PHS Amsterdam dataset was DCT notification, MCT notification, having symptoms without notification, or unknown: all individuals who reported having received a CoronaMelder notification as a reason for testing were included in the DCT group, individuals who reported having been contacted by the MCT but did not report a CoronaMelder notification were included in the MCT group, and individuals who reported having symptoms as a reason for testing and not DCT or MCT were included in the symptoms group. The hierarchy in the first RDT study was DCT notification, MCT notification, having been notified by an index case, testing at one’s own initiative (self), and unknown; symptoms were never a reason for testing because only asymptomatic individuals were eligible. The hierarchy in the second RDT study was DCT notification, MCT notification, having been notified by an index case, having a housemate who tested positive, testing at one’s own initiative (self), having received an unknown type of notification, having symptoms without notification, another reason for testing, and an unknown reason for testing. We used the last date of exposure/contact, and the date a test sample was taken, to calculate the exposure-testing interval in all datasets.

The adequacy of the Weibull models was checked for each dataset, and the log-likelihood test statistics showed that each model was statistically significantly better than the null-model (p <.001). The event time ratio (ETR), which signifies the relative difference in time intervals, the 95% confidence interval (CI), and the p-value were reported.

**SUPPLEMENTARY RESULTS**

In all three datasets, age and gender distributions differed statistically significantly across the reasons for testing groups, but these differences were small and considered not meaningful (Tables S1A-C). Among individuals testing because of a DCT or MCT notification in the first RDT study, the proportions testing in West Brabant were statistically significantly higher than the proportions in the overall group; this was the other way around for Rotterdam (Table S1B). In both regions, the MCT program was scaled down for approximately the first month of the two-month data collection period. Among individuals testing because of a MCT notification in the second RDT study, the proportion testing in Zwolle was statistically significantly higher, and in West-Brabant lower, than the proportions in the overall group (Table S1C). During this study period, the MCT program was only scaled down in West-Brabant and Rotterdam, and not in Zwolle.

Among people who had a positive SARS-CoV-2 PCR test, the median PCR cycle threshold (Ct) value as a proxy of SARS-CoV-2 viral load ranged from 23.4 to 27.2 in the first RDT study (Table S1B), and from 20.9 to 27.1 in the second RDT study, across reasons for testing (Table S1C). In both studies, the median Ct values were lowest in the DCT group and highest in the unknown reasons for testing groups, but these differences did not reach significance in the first RDT study. The second RDT dataset additionally included data on COVID-19 vaccination status and having had a previous SARS-CoV-2 infection. The proportions of both variables were differentially distributed across reasons for testing (reaching significance for the former only), but the numbers were small (Table S1C).

In the Weibull regression models of the PHS Amsterdam MCT subset, being aged 15-59 years was associated with a statistically significantly shorter exposure-testing interval than being in the youngest or oldest age groups, and male gender with a marginally longer interval (Table 2A). In the first RDT study, those aged 60 or older had a statistically significantly longer mean interval than those in the 16-29 years group, but age was not associated with interval in the second RDT study (Tables 2B-C). In both RDT studies, gender was not associated with interval in any of the time-to-event models.

The tobit regression models showed similar results as the Weibull models for the PHS Amsterdam MCT subset and the second RDT study (Tables S3A and S3C). For the first RDT study, the results of the Weibull and tobit models did show some differences (Table S3B), but the tobit model was considered not to be a good fit for this dataset because testing peaked on day five after exposure due to the testing policy for asymptomatic close contacts at that time.

**Figure S1:	CoronaMelder smartphone application**

| 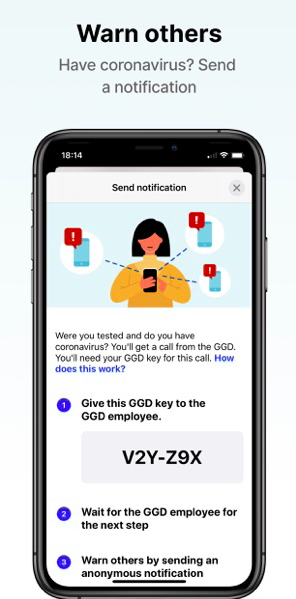 | 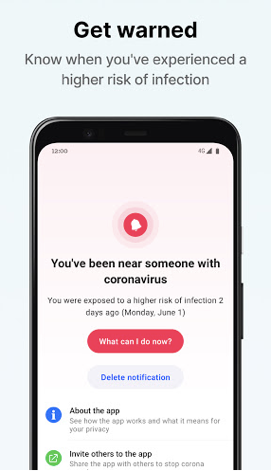 |
| --- | --- |
| Unique Public Health Service Key that would appear in the CoronaMelder app after the user tested SARS-CoV-2 positive | CoronaMelder exposure notification |

**Source:** De Winter et al, 2020. *Duidingsrapportage CoronaMelder: Informatiebeveiliging en privacybescherming* (report in Dutch language). Available at: https://www.rijksoverheid.nl/documenten/rapporten/ 2020/08/28/duidingsrapportage-coronamelder-informatiebeveiliging-en-privacybescherming-stand-van-zaken-lanceringsadvies (accessed Feb 5, 2023).

#### Figure S2: Participant flow diagram PHS Amsterdam datasets

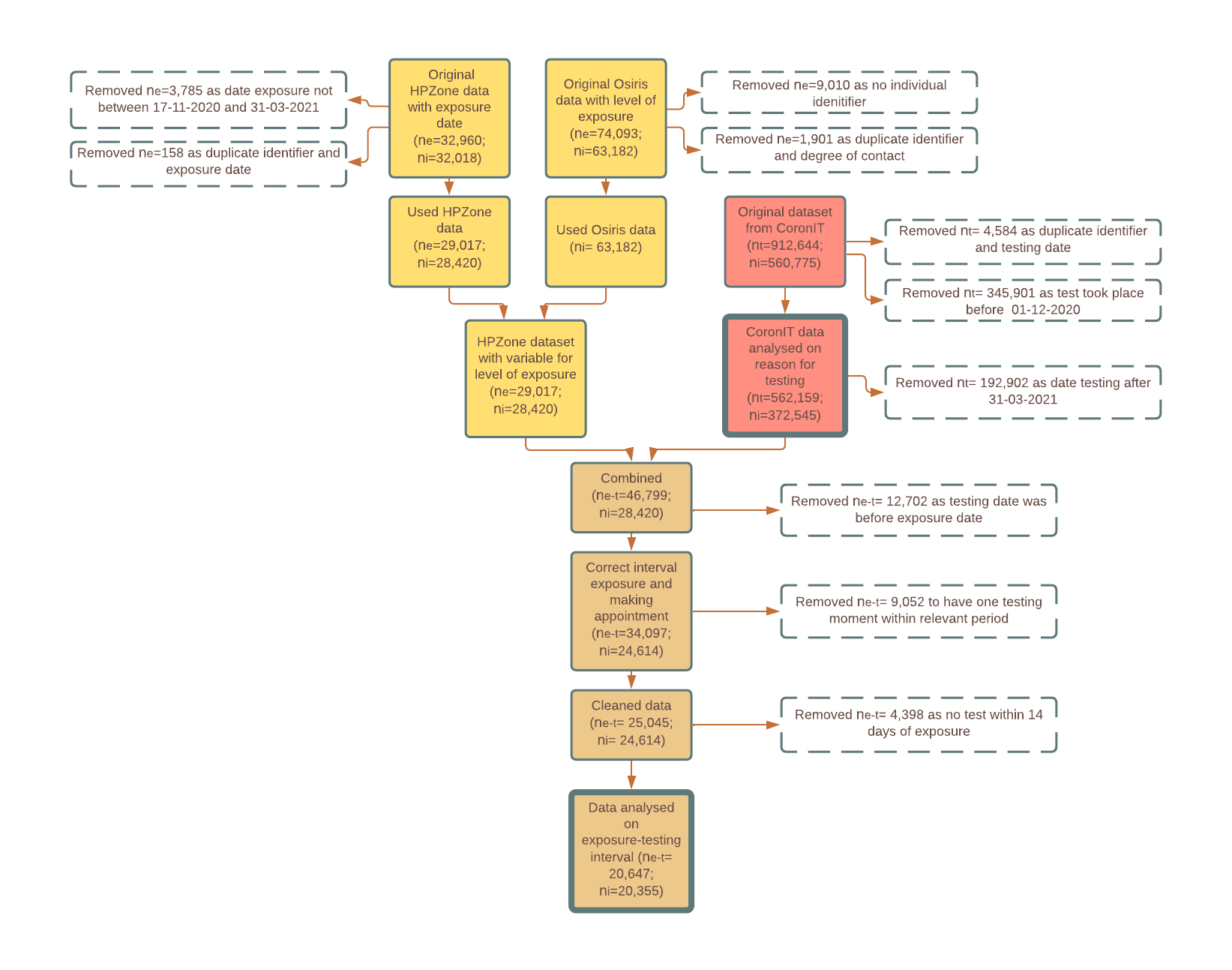
 Abbreviations: n_e_= exposure, n_t_= test, n_e-t_= interval exposure to testing, n_i_= individual

#### Figure S3: Questionnaires used in the first RDT and second RDT studies

| **A: First RDT study (asymptomatic individuals)^1^** | **B. Second RDT study^2^** |
| --- | --- |
| 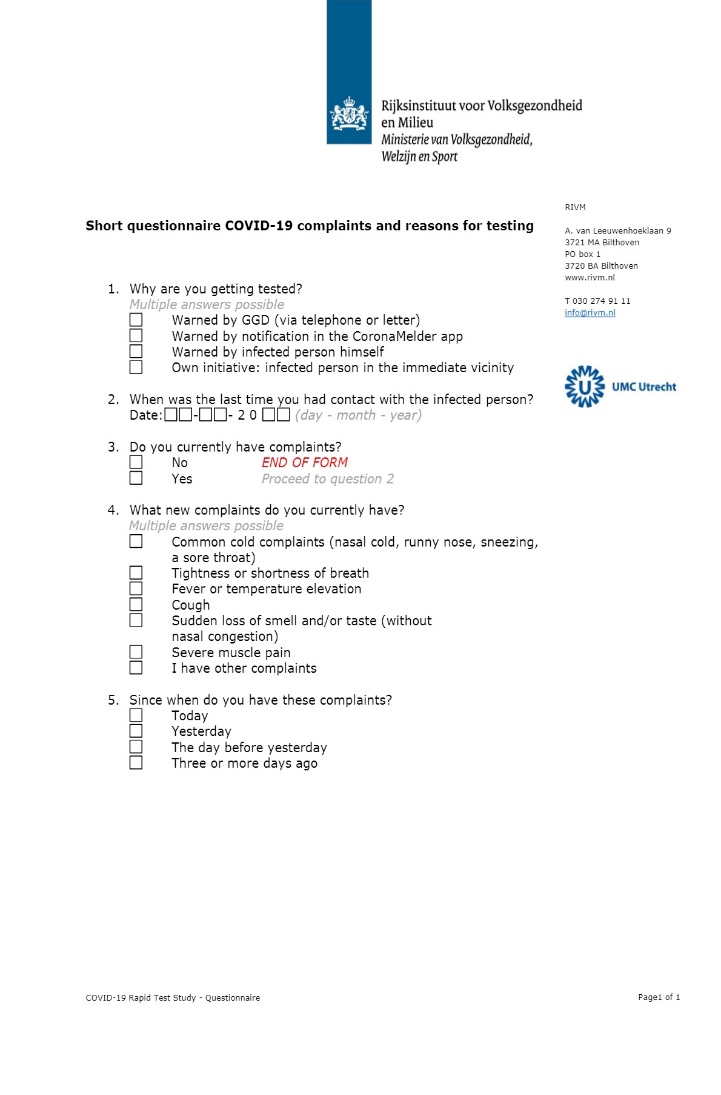 | 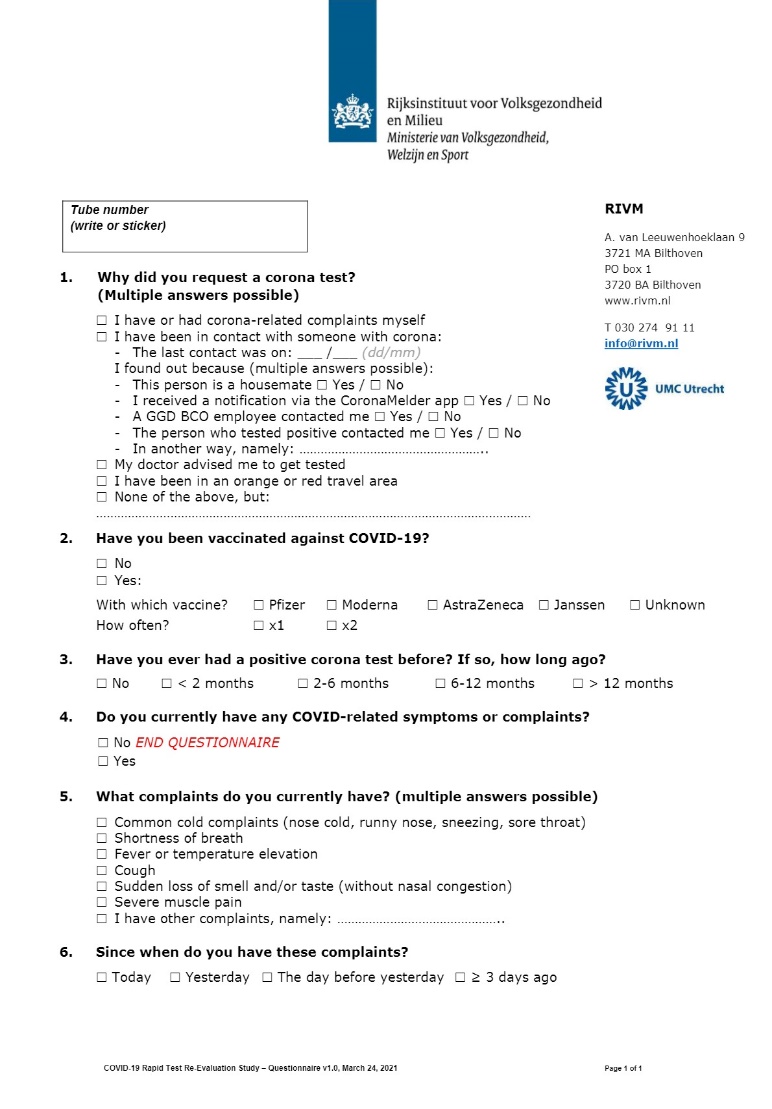 |

Abbreviations: RDT=rapid diagnostic test

1. The study period is 14 December 2020- 6 February 2021.
2. The study period is 12 April- 14 June 2021.

#### Figure S4: Participant flow diagrams first and second RDT studies

**A: First RDT study (asymptomatic close contacts)^1^**

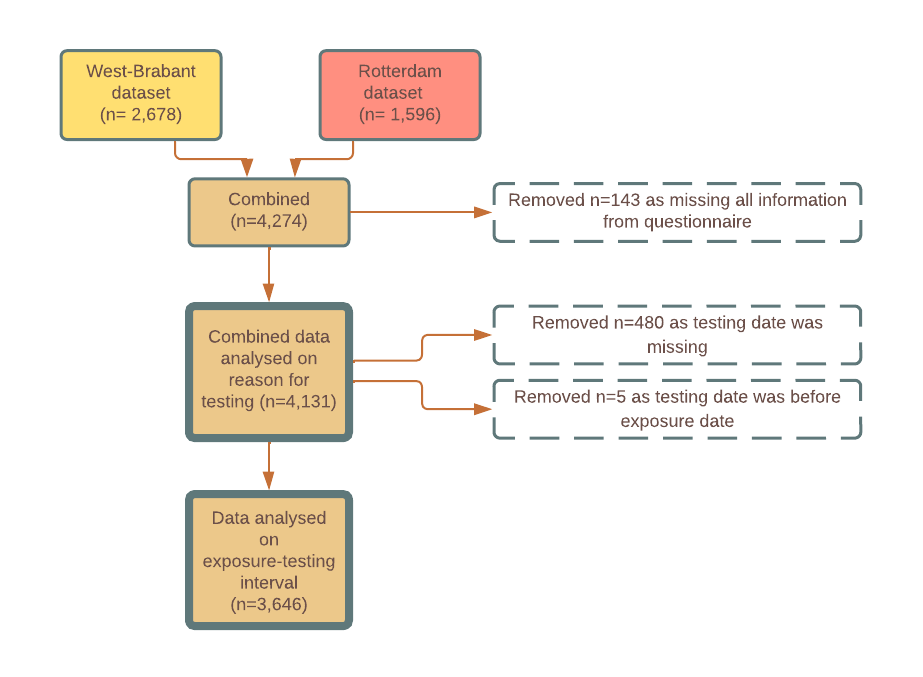

**B. Second RDT study^2^**

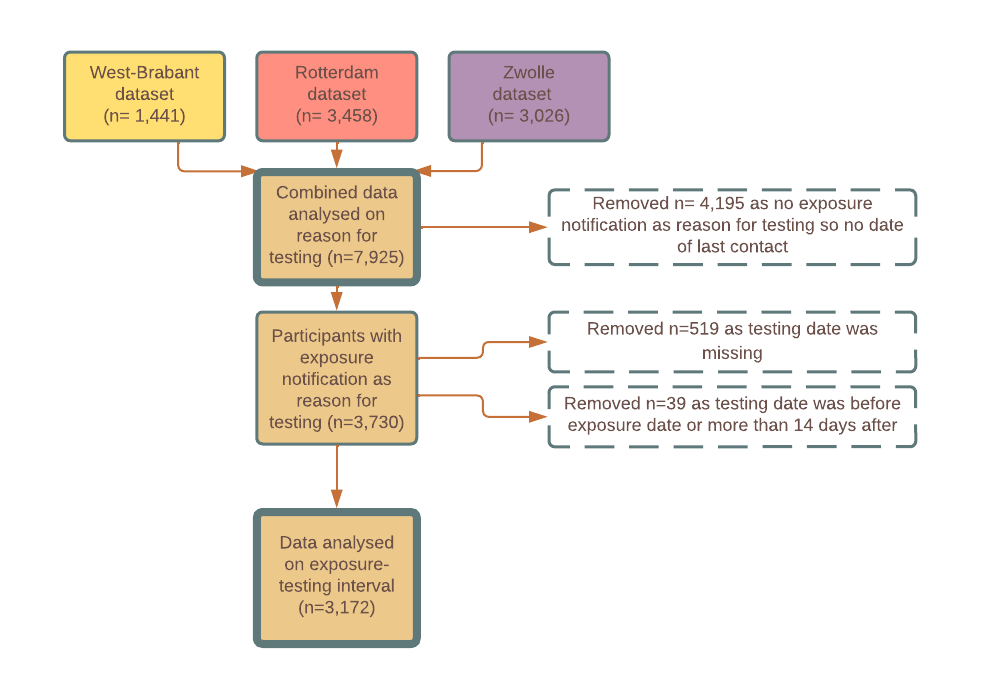

Abbreviations: RDT=rapid diagnostic test.

1. The study period was 14 December 2020 - 6 February 2021.
2. The study period was 12 April - 14 June 2021. Only participants who reported a close contact were asked the date of last exposure, and some dates were missing or incorrectly reported (e.g. before the testing date or more than 14 days after the testing date).

#### Table S1A: Test population characteristics by reason for testing – PHS Amsterdam

|  | **DCT**  n_t_= 8,231  (1.46%) | **MCT**  n_t_= 27,513  (4.89 %) | **Symptoms**  n_t_= 394,408  (70.16%) | **Unknown**  n_t_= 132,007  (23.48%) | **Total^1^**  n_t_= 562,159 | **p-value^2^** |
| --- | --- | --- | --- | --- | --- | --- |
| **Median age in years^3^** *(IQR)*  *[Range]* | 35  (27- 51)  [0-91] | 27  (12- 45)  [0-100] | 33  (24- 45)  [0-99] | 31  (21- 47)  [0-98] | 32  (23- 46)  [0-100] | <0.01 |
| **Gender^3^**  *Female,* n_t_ *(%)* | 4,467 (54.33) | 14,627 (53.25) | 218,795 (55.57) | 68,994 (52.44) | 306,883 (54.70) | <0.01 |
| **Municipality^3^**  *Amsterdam,* n_t_ *(%)*  *Aalsmeer,* n_t_ *(%)*  *Amstelveen,* n_t_ *(%)*  *Diemen,* n_t_ *(%)*  *Ouder-Amstel,* n_t_ *(%)*  *Uithorn,* n_t_ *(%)* | 6,782 (82.80)  232 (2.83)  604 (7.37)  224 (2.73)  127 (1.55)  222 (2.71) | 21,339 (77.77)  1,339 (4.88)  2,491 (9.08)  781 (2.85)  519 (1.89)  971 (3.54) | 329,544 (84.00)  9,912 (2.53)  28,238 (7.20)  10,204 (2.60)  5,167 (1.32)  9,232 (2.35) | 100,280 (76.25)  5,760 (4.38)  13,612 (10.35)  4,763 (3.62)  2,335 (1.78)  4,764 (3.62) | 457,945 (81.86)  17,243 (3.08)  44,945 (8.03)  15,972 (2.85)  8,148 (1.46)  15,189 (2.72) | <0.01 |
| **Symptoms**  *Yes,* n_t_ *(%)* | 2,940 (35.72) | 8,619 (31.33) | 394,408 (100) | 0 (0) | 405,967 (72.22) | <0.01 |
| **Test result^4^**  *Positive,* n_t_ *(%)* | 498 (6.09) | 4,821 (17.62) | 36,308 (9.26) | 9,895 (7.54) | 51,522 (9.22) | <0.01 |

Abbreviations: DCT=digital contact tracing; MCT=manual contact tracing; IQR=interquartile range

1. Includes 562,159 tests (n_t_) by 372,545 individuals (n_i_) between 1 December 2020 – 31 May 2021. The reason for testing categories are based on a hierarchy as explained in the methods. Missing values for test result (n_t_ = 3,480), age (n_t_ = 196), gender (n_t_ = 1,130), and municipality (n_t_ = 2,717).
2. Pearson’s Chi-squared for categorical variables and Kruskal-Wallis for continuous variables to determine differential distribution across reasons for testing. For further analysis in case of a statistically significant difference, see methods.
3. The statistically significant difference among the groups is not meaningful.
4. The test positivity percentage was statistically significantly lower among those testing after a DCT notification or for an unknown reason, and higher after a MCT notification.

#### Table S1B: Test population characteristics by reason for testing – first RDT study (asymptomatic close contacts)

|  | **DCT**  (n= 293, 7.09%) | **MCT**  (n= 507, 12.27%) | **Index**  (n= 2,409, 58.32%) | **Self**  (n= 479, 11.60%) | **Unknown**  (n= 443, 10.72%) | **Total^1^**  (n= 4,131) | **p^2^** |
| --- | --- | --- | --- | --- | --- | --- | --- |
| **Median age in years^3^** *(IQR)*  *[Range]* | 50  (35- 61)  [16-82] | 45  (28- 57)  [16-86] | 42  (27- 56)  [16-96] | 48  (32- 58)  [16-89] | 42  (27- 56)  [16-94] | 44  (28- 57)  [16-96] | <0.01 |
| **Gender**  *Female, n (%)* | 140 (47.95) | 245 (48.32) | 1,208 (50.38) | 219 (46.01) | 236 (53.39) | 2,048 (49.77) | 0.19 |
| **Test region^4^**  *West-Brabant*  *Rotterdam* | 206 (70.55)  86 (29.45) | 362 (71.40)  145 (28.60) | 1,286 (53.54)  1,116 (46.46) | 361 (75.68)  116 (24.32) | 328 (74.04)  115 (25.96) | 2,543 (61.71)  1,578 (38.29) | <0.01 |
| **Symptoms^5^**  *Yes, n (%)* | 13 (4.47) | 24 (4.75) | 205 (8.57) | 70 (14.71) | 65 (14.71) | 377 (9.18) | <0.01 |
| **Test result^6^**  *Positive, n (%)* | 10 (3.41) | 50 (9.86) | 196 (8.14) | 53 (11.06) | 40 (9.03) | 349 (8.45) | <0.01 |
| ***Median Ct rounds^,7^***  *(IQR)*  *Ct≤30, n (%)*  *Ct>30, n (%)* | 23.42  (21.91- 28.47)  8 (80.00)  2 (20.00) | 25.10  (20.22- 30.27)  37 (74.00)  13 (26.00) | 24.11  (20.74-32.24)  139 (70.92)  57 (29.08) | 25.10  (21.24-28.87)  41 (77.36)  12 (22.64) | 27.17  (22.60-32.20)  23 (57.50)  17 (42.50) | 24.90  (20.99-31.32)  248 (71.06)  101 (28.94) | 0.45 |

Abbreviations: Ct=Cycle threshold; DCT=digital contact tracing; Index=a person who tested SARS-CoV-2 positive; IQR=interquartile range; MCT=manual contact tracing; Self=testing at one’s own initiative.

1. Includes 4,131 tests by 4,131 participants between 14 December 2020- 6 February 2021. The reason for testing categories are based on a hierarchy as explained in the methods. Missing values for symptoms (n= 25), age (n= 10), gender (n= 16), and test location (n= 10).
2. Pearson’s Chi-squared for categorical variables and Kruskal-Wallis for continuous variables to determine differential distribution across reasons for testing. For further analysis in case of a statistically significant difference, see methods.
3. The statistically significant difference among the groups is not meaningful.
4. Among individuals testing because of an index notification, the proportion testing in West Brabant was statistically significantly lower as compared to the group average. This proportion was statistically significantly higher for all other reasons for testing. The reverse was true for Rotterdam.
5. Symptoms were present in a statistically significantly lower percentage of those testing after a DCT or MCT notification and a statistically significantly higher percentage in the Self and Unknown reason groups.
6. Test positivity was statistically significantly lower among those testing after a DCT notification.
7. Only the Ct-value of participants (n=349) with a positive test result were included (by definition, the Ct value is 45 in those testing negative). The Ct 30 cut-off is often used as a proxy of infectiousness.

#### Table S1C: Test population characteristics by reason for testing – second RDT study

|  | **DCT**  (n= 192;  2.42%) | **MCT**  (n= 152;  1.92%) | **Index**  (n= 1,495;  18.86%) | **Housemate**  (n= 880  11.10%) | **Self**  (n= 240  3.03%) | **Unknown notification**  (n= 771; 9.73%) | **Symptoms**  (n= 3,586; 45.25%) | **Other**  (n=315  3.97%) | **Unknown reason**  (n=294; 3.71%) | **Total^1^**  (n=7,925) | **p-value^2^** |
| --- | --- | --- | --- | --- | --- | --- | --- | --- | --- | --- | --- |
| **Median age in years^3^** *(IQR)*  *[Range]* | 40  (26- 51)  [18- 84] | 33  (24- 53)  [16-78] | 31  (24- 46)  [16-82] | 33  (23- 50)  [16-82] | 38  (26- 51)  [16-73] | 37  (25- 54)  [16-91] | 37  (28- 48)  [16-88] | 46  (31- 62)  [17-84] | 43  (29- 60)  [17-84] | 36  (26- 50)  [16-91] | <0.01 |
| **Gender^3^**  *Female,* n_t_ *(%)* | 101 (53.16) | 82 (54.30) | 756 (50.74) | 463 (52.67) | 138 (57.50) | 362 (47.07) | 1,914 (53.54) | 137 (43.49) | 143 (48.81) | 4,096 (51.83) | <0.01 |
| **Test region^4^**  *West-Brabant*  *Rotterdam*  *Zwolle* | 51 (26.56)  88 (45.83)  53 (27.60) | 8 (5.26)  43 (28.29)  101 (66.45) | 380 (25.42)  687 (45.95)  428 (28.63) | 211 (23.98)  436 (49.55)  233 (26.48) | 72 (30.00)  92 (38.33)  76 (31.67) | 230 (29.83)  405 (52.53)  136 (17.64) | 371 (10.35)  1464 (40.83)  1751 (48.83) | 55 (17.46)  128 (40.63)  132 (41.90) | 63 (21.43)  115 (39.12) 116 (39.46) | 1,441 (18.18)  3,458 (43.63)  3,026 (38.18) | <0.01 |
| **Symptoms^5^**  *Yes,* n *(%)* | 38 (19.79) | 43 (28.29) | 471 (31.59) | 287 (32.76) | 73 (30.80) | 242 (31.59) | 3,586 (100.0) | 130 (41.53) | 62 (55.36) | 4,932 (62.23) | <0.01 |
| **Test result^6^**  *Positive,* n *(%)* | 7 (3.65) | 11 (7.24) | 140 (9.36) | 201 (22.84) | 22 (9.17) | 105 (13.62) | 241 (6.72) | 17 (5.40) | 29 (9.86) | 773 (9.75) | <0.01 |
| ***Median Ct rounds^7^***  *(IQR)*  *Ct≤30, n (%)*  *Ct>30, n (%)* | 20.89  (20.05-24.07)  7 (100.00)  0 (0) | 22.00  (18.70-27.20)  9 (100.00)  0 (0) | 23.64  (20.79-29.57)  103 (76.87)  31 (23.13) | 23.14  (20.50-29.00)  161 (82.56)  32 (17.44) | 22.69  (19.15-28.53)  19 (86.36)  3 (13.64) | 22.80  (20.60-27.74)  84 (81.55)  19 (18.45) | 21.96  (19.70-25.85)  206 (88.79)  26 (11.21) | 27.13  (22.40-34.40)  11 (64.71)  6 (35.29) | 26.22  (22.22-31.02)  19 (67.86)  9 (32.14) | 22.95  (20.30-27.90)  619 (82.86)  128 (17.14) | <0.01 |
| **Vaccinated^3^**  *Yes,* n *(%)* | 34 (17.71) | 36 (23.68) | 128 (8.56) | 87 (9.91) | 32 (13.33) | 105 (13.64) | 414 (11.55) | 67 (21.27) | 37 (33.64) | 940 (12.15) | <0.01 |
| **Prior infection**  *Yes,* n *(%)* | 13 (6.77) | 17 (11.18) | 150 (10.06) | 72 (8.23) | 14 (5.86) | 68 (8.91) | 292 (8.16) | 36 (11.50) | 14 (12.61) | 676 (8.76) | 0.07 |

Abbreviations: Ct=Cycle threshold; DCT=digital contact tracing; Index=a person who tested SARS-CoV-2 positive; IQR=interquartile range; MCT=manual contact tracing; Self=testing at one’s own initiative.

1. Includes 7,925 tests by 7,925 participants between 12 April- 14 June 2021. The reason for testing categories are based on a hierarchy as explained in the methods. Missing values for symptoms (n=200), Ct rounds (n=26), age (n=19), gender (n=23), vaccination status (n=190), and prior SARS-CoV-2 infection (n= 210).
2. Pearson’s Chi-squared for categorical and Kruskal-Wallis for continuous variables to determine differential distribution across reasons for testing. For further analysis in case of a statistically significant difference, see methods.
3. The statistically significant difference among the groups is not meaningful.
4. Among individuals testing because of a MCT notification, the proportion testing in Zwolle was statistically significantly higher, and in West-Brabant lower. Among individuals testing because of an index notification, the proportion testing in Zwolle was statistically significantly lower and in West-Brabant higher. Among individuals testing because of a housemate or unknown notification, the proportion testing in Zwolle was statistically significantly lower and in Rotterdam and Brabant higher. Among individuals testing because of having symptoms, the proportion testing in Zwolle was statistically significantly higher and in Rotterdam and Brabant lower.
5. Includes either symptoms as reason for testing or presence of symptoms. The statistically significant difference among the groups is not meaningful due to the group testing based on symptoms.
6. Test positivity was statistically significantly higher after a notification from a housemate or an unknown notification, and lower among those testing based on symptoms.
7. Only the Ct-value of participants with a positive test result were included (by definition, the Ct value is 45 in those testing negative). The Ct 30 cut-off is often used as a proxy of infectiousness. The statistically significant difference among the groups is not meaningful.

#### Table S2A: Mean (SD) interval in days between first exposure and testing – PHS Amsterdam MCT subset

|  | **DCT**  (n_e-t_= 297;  1.44%) | **n_e-t_ included**  **(%)** | **MCT**  (n_e-t_= 6,624;  32.08%) | **n_e-t_ included**  **(%)** | **Symptoms**  (n_e-t_= 6,684;  32.37%) | **n_e-t_ included**  **(%)** | **Unknown**  (n_e-t_= 7,042; 34.11%) | **n_e-t_ included**  **(%)** | **Total^1^**  (n_e-t_=20,647) | **Total n_e-t_ included**  **(%)** |
| --- | --- | --- | --- | --- | --- | --- | --- | --- | --- | --- |
| **All:** *Mean (SD)* | 4.22 (2.71) |  | 4.49 (2.36) |  | 3.09 (2.71) |  | 4.11 (2.47) |  | 3.91 (2.59) |  |
| **Age in years:** *0-14*  *15-29*  *30-44*  *45-59*  *60+* | 5.22 (4.29)  4.12 (2.19)  4.04 (2.76)  4.13 (2.97)  4.65 (2.78) | 9 (3)  90 (30)  77 (26)  75 (25)  46 (15) | 4.54 (2.52)  4.54 (2.21)  4.41 (2.41)  4.42 (2.34)  4.57 (2.23) | 1,801 (27)  1,601 (24)  1,232 (19)  1,398 (21)  592 (9) | 3.42 (2.91)  3.06 (2.53)  2.96 (2.71)  2.98 (2.92)  3.35 (2.71) | 840 (13)  2,616 (39)  1,400 (21)  1,308 (20)  520 (8) | 4.42 (2.39)  3.93 (2.40)  4.04 (2.54)  3.87 (2.53)  4.37 (2.54) | 1,870 (27)  1,941 (28)  1,163 (17)  1,346 (19)  722 (10) | 4.29 (2.58)  3.72 (2.48)  3.77 (2.65)  3.78 (2.67)  4.16 (2.55) | 4,520 (22)  6,248 (30)  3,872 (19)  4,127 (20)  1,880 (9) |
| **Gender:** *Female*  *Male* | 4.17 (2.62)  4.28 (2.83) | 162 (55) 135 (45) | 4.44 (2.35)  4.55 (2.38) | 3,487 (53)  3,125 (47) | 3.05 (2.65)  3.13 (2.79) | 3,694 (55)  2,977 (45) | 4.11 (2.43)  4.13 (2.51) | 3,587 (51)  3,433 (49) | 3.86 (2.55)  3.96 (2.63) | 10,930 (53)  9,670 (47) |
| **Municipality:**  *Amsterdam*  *Aalsmeer*  *Amstelveen*  *Diemen*  *Ouder-Amstel*  *Uithorn* | 4.28 (2.75)  4.76 (2.19)  3.66 (2.33)  2.50 (1.52)  4.38 (3.54)  4.42 (3.13) | 218 (73)  17 (6)  29 (10)  6 (2)  8 (3)  19 (6) | 4.54 (2.38)  4.61 (2.17)  4.15 (2.25)  4.84 (2.40)  3.95 (2.16)  4.31 (2.53) | 5,091 (77) 323 (5)  635 (10)  220 (3)  129 (2)  226 (3) | 3.12 (2.72)  2.43 (2.46)  3.02 (2.64)  3.02 (2.65)  3.51 (2.92)  2.83 (2.95) | 5,497 (82)  181 (3)  528 (8)  196 (3)  107 (2)  175 (3) | 4.14 (2.45)  3.71 (2.62)  4.18 (2.44)  3.88 (2.55)  3.90 (2.12)  4.28 (2.82) | 5,461 (78)  291 (4)  680 (10)  246 (3)  129 (2)  235 (3) | 3.92 (2.6)  3.81 (2.54)  3.83(2.49)  3.93(2.63)  3.82(2.42)3.91(2.83) | 16,267 (79)  812 (4)  1,872 (9)  668 (3)  373 (2)  655 (3) |
| **Type of contact:^2^**  *Household*  *Close, long*  *Close, short*  *Other contact*  *Case* | 4.41 (3.41)  4.78 (1.59)  4.67 (1.37)  3.00 (1.41)  3.33 (2.61) | 109 (37) 93 (32)  6 (2)  2 (1)  85 (29) | 4.54 (2.58)  5.16 (1.51)  5.22 (1.51)  5.29 (0.99)  3.59 (2.51) | 2,982 (45)  1,920 (29)  68 (1)  14 (0)  1,622 (25) | 2.78 (2.93)  3.89 (2.21)  3.90 (1.96)  2.69 (2.02)  2.70 (2.75) | 2,254 (34)  1,939 (29)  72 (1)  13 (0)  2,382 (36) | 3.88 (2.74)  4.88 (1.70)  5.35 (1.36)  4.85 (1.82)  3.18 (2.67) | 2,825 (40)  2,583 (37)  89 (1)  13 (0)  1,507 (21) | 3.83 (2.84)  4.67 (1.89) 4.85 (1.72)  4.24 (1.96)  3.09 (2.68) | 8,170 (40)  6,535 (32)  235 (1)  42 (0)  5,596 (27) |
| **Symptoms:** *No*  *Yes* | 4.98 (2.53)  3.40 (2.67) | 154 (52)  143 (48) | 4.85 (2.15)  3.55 (2.62) | 4,796 (72)  1,828 (28) | ---  3.09 (2.71) | 0 (0)  6,684 (100) | 4.11 (2.47)  --- | 7,042 (100)  0 (0) | 4.42 (2.38)  3.19 (2.70) | 11,992 (58)  8,655 (42) |
| **Test result:** *Negative*  *Positive* | 4.45 (2.74)  3.55 (2.53) | 222 (75)  75 (25) | 4.72 (2.28)  3.71 (2.49) | 5,104 (77)  1,493 (23) | 3.17 (2.68)  2.92 (2.79) | 4,552 (68)  2,107 (32) | 4.23 (2.41)  3.54 (2.66) | 5,790 (83)  1,226 (17) | 4.09 (2.53)  3.33 (2.69) | 15,668 (76)  4,901 (24) |

Abbreviations: DCT=digital contact tracing; MCT=manual contact tracing; SD= standard deviation

1. Includes 20,647 exposure- testing intervals (n_e-t_) by 20,355 individuals (n_i_) between 1 December 2020- 31 March 2021. Missing values for type of contact (n_e-t_= 69), test result (n_e-t_= 78), and gender (n_e-t_= 47).
2. “Close” is defined as within 1.5 meters of an infectious person; “long” as more than 15 minutes; “short” as 15 minutes or less but with high intensity (e.g. coughing in someone’s face, kissing); “household” as a close contact within the same residence; and “other” as any other contact with an infectious person.

#### Table S2B: Mean (SD) interval in days between last exposure and testing – first RDT study (asymptomatic close contacts)

|  | **DCT**  (n= 285;  7.82%) | **n_e-t_ included**  **(%)** | **MCT**  (n= 503;  13.80%) | **n_e-t_ included**  **(%)** | **Index**  (n= 2,389;  65.52%) | **n_e-t_ included**  **(%)** | **Self**  (n= 444;  12.18% | **n_e-t_ included**  **(%)** | **Unknown**  (n= 25;  0.69%) | **n_e-t_ included**  **(%)** | **Total^1^**  (n= 3,646) | **n_e-t_ included**  **(%)** |
| --- | --- | --- | --- | --- | --- | --- | --- | --- | --- | --- | --- | --- |
| **All**  *Mean (SD)* | 5.17 (1.39) |  | 5.09 (1.44) |  | 4.96 (1.34) |  | 4.49 (2.20) |  | 4.16 (2.67) |  | 4.93 (1.51) |  |
| **Age in years**  *16-29*  *30-44*  *45-59*  *60+* | 5.14 (1.54)  5.15 (0.90)  5.02 (1.31)  5.39 (1.75) | 42 (15)  71 (25)  95 (33)  76 (27) | 5.12 (1.35)  5.00 (1.33)  5.07 (1.45)  5.17 (1.68) | 135 (27)  114 (23)  151 (30)  103 (20) | 4.91 (1.25)  5.10 (1.36)  4.83 (1.38)  5.06 (1.37) | 704 (30)  565 (24)  672 (28)  441 (19) | 4.87 (2.03)  4.29 (2.18)  4.51 (2.36)  4.27 (2.13) | 98 (22)  91 (21)  160 (36)  94 (21) | 4.29 (3.30)  4.33 (1.15)  4.60 (2.88)  3.80 (2.74) | 7 (28)  3 (12)  5 (20)  10 (40) | 4.94 (1.39)  5.00 (1.46)  4.83 (1.58)  4.99 (1.63) | 986 (27)  844 (23)  1,083 (30)  724 (20) |
| **Gender**  *Female*  *Male* | 5.31 (1.22)  5.05 (1.54) | 137 (48)  147 (52) | 5.12 (1.39)  5.05 (1.49) | 245 (49)  258 (51) | 4.99 (1.27)  4.93 (1.41) | 1,194 (50)  1,184 (50) | 4.41 (2.26)  4.55 (2.16) | 204 (46)  238 (54) | 3.56 (2.79)  4.50 (2.63) | 9 (36)  16 (64) | 4.96 (1.46)  4.90 (1.57) | 1,789 (49)  1,843 (51) |
| **Test location**  *West-Brabant*  *Rotterdam* | 5.17 (1.58)  5.19 (0.82) | 199 (70)  85 (30) | 5.15 (1.54)  4.92 (1.14) | 358 (71)  145 (29) | 4.94 (1.49)  4.98 (1.15) | 1,272 (53)  1,110 (47) | 4.38 (2.39)  4.85 (1.47) | 336 (76)  107 (24) | 4.59 (2.50)  1.00 (1.73) | 22 (88)  3 (12) | 4.90 (1.70)  4.97 (1.17) | 2,187 (60)  1,450 (40) |
| **Symptoms**  *No*  *Yes* | 5.19 (1.42)  4.85 (0.90) | 270 (95)  13 (5) | 5.09 (1.46)  4.91 (1.12) | 479 (95)  23 (5) | 5.00 (1.25)  4.58 (2.01) | 2,174 (92)  199 (8) | 4.51 (2.12)  4.35 (2.73) | 388 (88)  55 (12) | 4.71 (2.37)  1.67 (2.89) | 21 (88)  3 (12) | 4.97 (1.44)  4.54 (2.10) | 3,332 (92)  293 (8) |
| **Test result**  *Negative*  *Positive* | 5.15 (1.41)  5.70 (0.95) | 275 (96)  10 (4) | 5.13 (1.29)  4.70 (2.39) | 453 (90)  50 (10) | 4.99 (1.30)  4.57 (1.72) | 2,195 (92)  194 (8) | 4.53 (2.18)  4.16 (2.37) | 395 (89)  49 (11) | 4.13 (2.78)  4.50 (0.71) | 23 (92)  2 (8) | 4.96 (1.46)  4.56 (1.95) | 3,341 (92)  305 (8) |

Abbreviations: DCT=digital contact tracing; Index=a person who tested SARS-CoV-2 positive; MCT=manual contact tracing; SD=standard deviation; Self=testing at one’s own initiative.

1. Includes 3,646 exposure-test intervals in 3,646 participants between 14 December 2020- 6 February 2021. Only participants who reported a close contact were asked the date of last exposure and dates are missing (n=480) or not logical (before testing date or more than 14 days after testing, n=5). Additional missing values for symptoms (n=21), age (n=9), gender (n=14), and test location (n=9).

#### Table S2C: Mean (SD) interval in days between last exposure and testing – second RDT study

|  | **DCT**  (n= *153;*  *4.82*%) | **n**  **(%)** | **MCT**  (n= 139;  4.38%) | **n**  **(%)** | **Index** (n=1,419;  44.74%) | **n**  **(%)** | **Housemate**  (n=795;  25.06%) | **n**  **(%)** | **Self**  (n=147;  4.63%) | **n**  **(%)** | **Unknown** (n=519; 16.36%) | **n**  **(%)** | **Total^1^**  (n=3,172) | **Total n**  **(%)** |
| --- | --- | --- | --- | --- | --- | --- | --- | --- | --- | --- | --- | --- | --- | --- |
| **All**  *Mean (SD)* | 4.84 (1.59) |  | 5.11 (1.68) |  | 4.70 (1.52) |  | 3.12 (2.53) |  | 4.94 (1.90) |  | 4.47 (1.93) |  | 4.30 (2.04) |  |
| **Symptoms**  *No*  *Yes* | 4.74 (1.43)  5.22 (2.09) | 121 (79)  32 (21) | 4.94 (1.10)  5.64 (2.78) | 106 (76)  33 (24) | 4.75 (1.42)  4.60 (1.73) | 970 (69)  445 (31) | 3.44 (2.49)  2.47 (2.49) | 528 (67)  263 (33) | 5.15 (1.69)  4.62 (2.21) | 93 (64)  52 (36) | 4.66 (1.85)  4.03 (2.04) | 361 (70)  155 (30) | 4.44 (1.89)  4.00 (2.30) | 2,179 (69)  980 (31) |
| **Test result**  *Negative*  *Positive* | 4.83 (1.62)  5.14 (0.90) | 146 (95)  7 (5) | 5.06 (1.64)  5.88 (2.10) | 131 (94)  8 (6) | 4.72 (1.49)  4.53 (1.84) | 1,284 (90)  135 (10) | 3.29 (2.52)  2.54 (2.51) | 615 (77)  180 (23) | 4.95 (1.81)  4.87 (2.64) | 132 (90)  15 (10) | 4.63 (1.84)  3.39 (2.22) | 455 (88)  64 (12) | 4.42 (1.94)  3.52 (2.44) | 2,763 (87)  409 (13) |
| **Age in years**  *16-29*  *30-44*  *45-59*  *60+* | 4.81 (1.66)  4.75 (1.38)  5.03 (1.93)  4.82 (1.13) | 57 (37)  44 (29)  35 (23)  17 (11) | 4.84 (1.13)  6.29 (2.54)  4.35 (1.58)  5.17 (0.96) | 64 (46)  28 (20)  23 (17)  24 (17) | 4.58 (1.47)  4.87 (1.58)  4.76 (1.64)  4.71 (1.39) | 678 (48)  365 (26)  238 (17)  133 (9) | 3.52 (2.30)  2.29 (2.80)  3.13 (2.58)  2.64 (2.53) | 378 (48)  159 (20)  215 (27)  42 (5) | 5.16 (1.91)  4.89 (1.81)  4.59 (1.80)  5.09 (2.21) | 50 (34)  37 (25)  37 (25)  23 (16) | 4.52 (1.91)  4.16 (1.99)  4.51 (1.90)  5.04 (1.83) | 224 (43)  135 (26)  111 (21)  49 (9) | 4.34 (1.87)  4.26 (2.27)  4.18 (2.18)  4.54 (1.88) | 1,451 (46)  768 (24)  659 (21)  288 (9) |
| **Gender**  *Female*  *Male* | 4.96 (1.71)  4.70 (1.48) | 81 (54)  70 (46) | 5.09 (1.56)  5.13 (1.83) | 78 (56)  61 (44) | 4.77 (1.44)  4.63 (1.60) | 725 (51)  689 (49) | 3.12 (2.54)  3.13 (2.53) | 419 (53)  375 (47) | 4.93 (1.79)  4.95 (2.08) | 90 (61)  57 (39) | 4.65 (1.80)  4.32 (2.04) | 244 (47)  274 (53) | 4.36 (2.01)  4.24 (2.07) | 1,637 (52)  1,526 (48) |
| **Testing region**  *West-Brabant*  *Rotterdam*  *Zwolle* | 5.16 (1.00)  4.60 (1.92)  4.90 (1.48) | 43 (28)  68 (44)  42 (27) | 5.71 (2.93)  4.98 (1.87)  5.12 (1.46) | 7 (5)  42 (30)  90 (65) | 4.90 (1.13)  4.69 (1.68)  4.54 (1.55) | 362 (26)  651 (46)  406 (29) | 3.46 (2.64)  3.19 (2.47)  2.69 (2.49) | 196 (25)  384 (48)  215 (27) | 5.17 (1.22)  5.27 (2.19)  4.11 (1.89) | 47(32)  62 (42)  38 (26) | 4.60 (1.59)  4.50 (1.94)  4.19 (2.35) | 164 (32)  252 (49)  103 (20) | 4.53 (1.82)  4.29 (2.11)  4.11 (2.09) | 819 (26)  1,459 (46)  894 (28) |
| **Vaccinated**  *No*  *Yes* | 4.84 (1.57)  4.86 (1.75) | 131 (86)  22 (14) | 5.07 (1.83)  5.22 (1.01) | 107 (77)  32 (23) | 4.69 (1.52)  4.77 (1.57) | 1,303 (92)  116 (8) | 3.09 (2.51)  3.35 (2.77) | 716 (90)  78 (10) | 4.97 (1.97)  4.72 (1.36) | 129 (88)  18 (12) | 4.41 (1.92)  5.00 (1.99) | 461 (89)  58 (11) | 4.28 (2.03)  4.52 (2.06) | 2,847 (90)  324 (10) |
| **Prior infection**  *No*  *Yes* | 4.82 (1.59)  5.09 (1.64) | 142 (93)  11 (7) | 5.08 (1.53)  5.33 (2.66) | 124 (89)  15 (11) | 4.71 (1.54)  4.60 (1.40) | 1,272 (90)  143 (10) | 3.09 (2.55)  3.34 (2.18) | 725 (92)  67 (8) | 5.07 (1.83)  2.62 (1.77) | 138 (95)  8 (5) | 4.43 (1.93)  4.90 (1.93) | 470 (91)  48 (9) | 4.29 (2.05)  4.36 (1.90) | 2,871 (91)  292 (9) |

Abbreviations: DCT=digital contact tracing; Index=a person who tested SARS-CoV-2 positive; MCT=manual contact tracing; SD=standard deviation; Self=testing at one’s own initiative.

1. Includes 3,172 exposure-test intervals in 3,172 participants between 12 April- 14 June 2021. Only participants who reported a close contact were asked the date of last exposure and dates are missing (n=519) or not logical (before testing date or more than 14 days after testing, n=39). Additional missing values for symptoms (n=13), age (n= 6), gender (n=9), vaccination status (n=1), and prior SARS-CoV-2 infection (n= 9).

### Table S3A: Tobit regression model for exposure-test intervals – PHS Amsterdam MCT subset

|  | **Univariable analysis^1^ (n_e-t_= 20,647)** | | | **Multivariable analysis^1^ (n_e-t_= 20,647)** | | |
| --- | --- | --- | --- | --- | --- | --- |
|  | **Coefficient^2^** | **95% CI** | **p-value** | **Coefficient^2^** | **95% CI** | **p-value** |
| **Age in years:** *0-14*  *15-29*  *30-44*  *45-59*  *60+* | 0.60  *reference*  0.01  0.03  0.45 | 0.49-0.71  ---  -0.11-0.12  -0.09-0.15  0.30-0.61 | <0.01  ---  0.92  0.61  <0.01 | 0.32  *reference*  0.01  0.11  0.34 | 0.21-0.43  ---  -0.10-0.12  0.00-0.22  0.19-0.48 | <0.01  ---  0.85  0.05  <0.01 |
| **Gender:** *Female*  *Male* | *reference*  0.11 | ---  0.03-0.19 | ---  <0.01 | *reference*  0.07 | ---  -0.01-0.15 | ---  0.07 |
| **Municipality:**  Amsterdam  Surrounding area | *reference*  -0.09 | ---  -0.18-0.01 | ---  0.09 | *reference*  -0.13 | ---  -0.23-(-) 0.04 | ---  <0.01 |
| **Type of contact:^3^**  *Household*  *Close, long*  *Close, short*  *Other contact*  *Case* | *reference*  1.01  1.22  0.56  -0.84 | ---  0.92-1.10  0.85-1.59  -0.31-1.42  -0.94-(-)0.74 | ---  <0.01  <0.01  0.21  <0.01 | *reference*  1.00  1.22  0.61  -0.57 | *---*  0.91-1.09  0.86-1.58  -0.24-1.45  -0.66-(-)0.47 | ---  <0.01  <0.01  0.16  <0.01 |
| **DCT:** *No*  *Yes* | *reference*  0.36 | ---  0.03-0.70 | ---  0.03 | *reference*  0.48 | ---  0.15-0.80 | ---  <0.01 |
| **Symptoms***: No*  *Yes* | *reference*  -1.39 | ---  -1.47-(-)1.31 | ---  <0.01 | *reference*  -1.17 | ---  -1.25-(-)1.09 | ---  <0.01 |
| **Test result:** *Negative*  *Positive* | *Reference*  -0.89 | ---  -0.99-(-)0.80 | ---  <0.01 | *Not included* | --- | --- |

Abbreviations: CI=confidence interval; DCT=digital contact tracing; MCT=manual contact tracing.

1. Based on 20,647 exposure- testing intervals (n_e-t_) by 20,355 individuals (n_i_) between 1 December 2020- 31 March 2021. Missing values for type of contact (n_e-t_= 69), test result (n_e-t_= 78), and gender (n_e-t_= 47).
2. The Tobit coefficient represents the change in exposure-test interval (in days) for each unit change of the dependent variable.
3. “Close” is defined as within 1.5 meters of an infectious person; “long” as more than 15 minutes; “short” as 15 minutes or less but with high intensity (e.g. coughing in someone’s face, kissing); “household” as a close contact within the same residence; and “other” as any other contact with an infectious person.

#### Table S3B: Tobit regression model for the exposure-test intervals – first RDT study

|  | **Univariable analysis^1^ (n= 3,646)** | | | **Multivariable analysis^1^ (n= 3,646)** | | |
| --- | --- | --- | --- | --- | --- | --- |
|  | **Coefficient^2^** | **95% CI** | **p-value** | **Coefficient^2^** | **95% CI** | **p-value** |
| **Age in years***: 16-29*  *30-44*  *45-59*  *60+* | *reference*  0.06  -0.11  0.05 | *---*  -0.08-0.21  -0.25- 0.02  -0.10-0.20 | ---  0.40  0.11  0.53 | *reference*  0.04  -0.10  0.06 | ---  -0.10-0.19  -0.24-0.03  -0.09-0.21 | ---  0.54  0.14  0.46 |
| **Gender:** *Female*  *Male* | *reference*  -0.06 | ---  -0.16-0.05 | ---  0.28 | *reference*  -0.05 | ---  -0.15-0.05 | ---  0.32 |
| **Testing region:** *Brabant*  *Rotterdam* | *reference*  *0.08* | ---  -0.02-0.19 | ---  0.12 | *reference*  0.05 | ---  -0.06-0.15 | ---  0.37 |
| **DCT:** *No*  *Yes* | *reference*  0.27 | ---  0.08-0.46 | ---  <0.01 | *reference*  0.08 | *---*  -0.16-0.32 | ---  0.51 |
| **MCT:** *No*  *Yes* | *reference*  0.18 | ---  0.04-0.33 | ---  0.01 | *reference*  0.04 | ---  -0.14-0.22 | ---  0.64 |
| **Index:** *No*  *Yes* | *reference*  0.15 | ---  0.04-0.27 | ---  <0.01 | *reference*  -0.14 | ---  -0.34-0.07 | ---  0.19 |
| **Self:** *No*  *Yes* | *reference*  -0.56 | ---  -0.71-(-)0.41 | ---  <0.01 | *reference*  -0.63 | ---  -0.87-0.39 | ---  <0.01 |
| **Unknown** *No*  **Contact:** *Yes* | *reference*  -0.88 | ---  -1.49- (-)0.26 | ---  <0.01 | *reference*  -0.83 | ---  -1.49-(-)0.18 | ---  0.01 |
| **Symptoms:** *No*  *Yes* | *reference*  -0.45 | ---  -0.63-(-)0.26 | ---  <0.01 | *reference*  -0.36 | ---  -0.55-(-)0.17 | ---  <0.01 |
| **Test result***: Negative*  *Positive* | *reference*  -0.42 | *---*  -0.61-(-)0.24 | ---  <0.01 | *reference*  -0.36 | ---  -0.54-(-)0.18 | ---  <0.01 |

Abbreviations: CI= confidence interval; DCT=digital contact tracing; Index=a person who tested SARS-CoV-2 positive; MCT=manual contact tracing; Self=testing at one’s own initiative.

1. Includes 3,646 exposure-test intervals in 3,646 participants between 14 December 2020- 6 February 2021. Only participants who reported a close contact were asked the date of last exposure and dates are missing (n=480) or not logical (before testing date or more than 14 days after testing, n=5). Additional missing values for symptoms (n=21), age (n=9), gender (n=14), and test location (n=9).
2. The Tobit coefficient represents the change in exposure-test interval (in days) for each unit change of the dependent variable.

#### Table S3C: Tobit regression model for the exposure-test intervals – second RDT study

|  | **Univariable analysis^1^ (n=3,172)** | | | **Multivariable analysis^1^ (n=3,172)** | | |
| --- | --- | --- | --- | --- | --- | --- |
|  | **Coefficient^2^** | **95% CI** | **p-value** | **Coefficient^2^** | **95% CI** | **p-value** |
| **Age in years***: 16-29*  *30-44*  *45-59*  *60+* | *reference*  -0.13  -0.19  0.20 | ---  -0.32-0.06  -0.39-0.01  -0.08-0.48 | ---  0.17  0.06  0.16 | *reference*  -0.24  -0.11  -0.21 | ---  -0.41-(-)0.06  -0.30-0.08  -0.49-0.08 | ---  <0.01  0.27  0.16 |
| **Gender:** *Female*  *Male* | *reference*  -0.13 | ---  -0.28-0.03 | ---  0.11 | *reference*  -0.12 | ---  -0.26-0.02 | ---  0.10 |
| **Testing region:** *Brabant*  *Rotterdam*  *Zwolle* | *reference*  -0.24  -0.43 | ---  -0.42-(-)0.05  -0.64-(-)0.23 | ---  0.01  <0.01 | *reference*  -0.23  -0.55 | ---  -0.41-(-)0.06  -0.75-(-)0.35 | ---  <0.01  <0.01 |
| **DCT:** *No*  *Yes* | *reference*  0.38 | ---  0.07-0.69 | ---  0.02 | *reference*  0.41 | ---  0.10-0.71 | ---  <0.01 |
| **MCT:** *No*  *Yes* | *reference*  0.20 | ---  -0.07-0.46 | ---  0.15 | *reference*  0.67 | ---  0.41 0.92 | ---  <0.01 |
| **Index:** *No*  *Yes* | *reference*  0.52 | ---  0.37-0.68 | ---  <0.01 | *reference*  0.40 | ---  0.16-0.64 | ---  <0.01 |
| **Housemate:** *No*  *Yes* | *reference*  -1.78 | ---  -1.95-(-)1.62 | ---  <0.01 | *reference*  -1.67 | ---  -1.88-(-)1.46 | ---  <0.01 |
| **Self:** *No*  *Yes* | *reference*  0.70 | ---  0.34-1.06 | ---  <0.01 | *reference*  0.64 | ---  0.22-1.05 | ---  <0.01 |
| **Unknown** *No*  **Contact:** *Yes* | *reference*  0.22 | ---  0.01-0.42 | ---  0.04 | *reference*  0.11 | ---  -0.20-0.41 | ---  0.49 |
| **Symptoms:** *No*  *Yes* | *reference*  -0.50 | ---  -0.66-(-)0.33 | ---  <0.01 | *reference*  -0.32 | ---  -0.48-(-)0.16 | ---  <0.01 |
| **Test result***: Negative*  *Positive* | *reference*  -1.02 | ---  -1.24-(-)0.79 | ---  <0.01 | *reference*  -0.52 | ---  -0.74-(-)0.29 | ---  <0.01 |
| **Vaccinated:** *No*  *Yes* | *reference*  0.24 | ---  -0.01-0.49 | ---  0.06 | *reference*  0.19 | ---  -0.07-0.45 | ---  0.15 |
| **Previous** *No*  **infection:** *Yes* | *reference*  0.09 | ---  -0.17-0.36 | ---  0.49 | *reference*  0.02 | ---  -0.23-0.26 | ---  0.88 |

Abbreviations: CI= confidence interval; DCT=digital contact tracing; Index=a person who tested SARS-CoV-2 positive; MCT=manual contact tracing; Self=testing at one’s own initiative.

1. Includes 3,172 exposure-test intervals in 3,172 participants between 12 April- 14 June 2021. Only participants who reported a close contact were asked the date of last exposure and dates are missing (n=519) or not logical (before testing date or more than 14 days after testing, n=39). Additional missing values for symptoms (n=13), age (n= 6), gender (n=9), vaccination status (n=1), and prior infection (n= 9).
2. The Tobit coefficient represents the change in exposure-test interval (in days) for each unit increase of the dependent variable.
